# Recent cross-sectional prevalence studies in sub-Saharan Africa for communicable, maternal, neonatal, and nutritional diseases and conditions: a scoping review

**DOI:** 10.1101/2023.12.24.23300511

**Authors:** S Dolley, C Miller, P Quach, T Norman

**Affiliations:** Open Global Health; ConfluenceStat Inc.; OneShot Data, LLC; The Bill & Melinda Gates Foundation

**Keywords:** prevalence disease burden cross-sectional scoping review

## Abstract

**Background:** Cross-sectional prevalence studies provide benefits to policymakers, epidemiologists, trialists, and the future health of target and general populations. Too few of these studies are performed in hotspots of traditional global health disease burden. This results in a lack of recent, local, accurate prevalence estimates to inform policy, epidemiology, and the design of interventional randomized controlled trials that may be conducted in these regions.

**Objective:** This scoping review aims to establish a novel dataset usable as an observational baseline. The topic being analyzed is the set of characteristics describing recently published prospective cross-sectional prevalence studies in sub-Saharan Africa for humans affected with communicable, maternal, neonatal, and nutritional diseases and conditions.

**Methods:** This scoping review conducted a systematic literature search of PubMed. The search identified publications in the last four years describing completed cross-sectional prevalence studies in sub-Saharan Africa. Title and abstract screening was completed. Data extraction was performed on a random sample of the final dataset.

**Results:** This scoping review identified 868 titles and abstracts through our systematic search. Through our screening process, 394 of these were candidates eligible for inclusion in our dataset. Ultimately, 363 were in the final dataset. Of the 38 studies in the random sample, this scoping review found a large portion of the studies completed with no funding. Malaria was the predominant disease studied, followed by parasitic intestinal infection and malnutrition. Studies with funding were slower, from data collection to submission to a journal, than studies with no funding. Studies that use a national ethical review process tend to take longer than those using hospital or university institutional review boards.

**Conclusions:** Cross-sectional prevalence studies are happening in sub-Saharan Africa for many communicable, maternal, neonatal, and nutritional diseases and conditions. This healthy research ecosystem is filled with variety. A variety of data collection methods, sources of funding, types of study sites, and target populations exist. Many studies are self-funded by the principal investigator. These studies are rarely conducted for the explicit purpose of informing the designs of future randomized trials. Some trends might be observable in the data that point to causal factors for study speed or sample size.

## Introduction

### Cross-Sectional Prevalence Studies

Cross-sectional study design is “a type of observational study design…in a cross-sectional study, the investigator measures the outcome and the exposures in the participants at the same time. (Setia)”. A cross-sectional study to determine the prevalence of disease in humans would involve going out to directly measure participants at a single point in time. However, a retrospective design would involve mining historical clinical data to calculate estimations about populations. Table 1 lists definitions of types of cross-sectional studies considered in this review.

**Table 1.** Contemporary definitions of cross-sectional prevalence studies.

| Study type | Study type definition |
| --- | --- |
| Cross-sectional study | Cross-sectional study: “a type of observational study design. In a cross-sectional study, the investigator measures the outcome and the exposures in the participants at the same time (Setia)”. |
| Cross-sectional prevalence study | Cross-sectional prevalence study: An observational, analytical study that measures a target population to identify disease burden or rate of a condition at a single point in time. |
| Retrospective cross-sectional prevalence study | Retrospective cross-sectional prevalence study: a point-in-time assessment, using historical or secondary use data in a systematic and rigorous way, for estimations about prevalence of a disease in population(s) at a prior single point in time. |
| Prospective cross-sectional prevalence study | Prospective cross-sectional prevalence study: a non-retrospective, point-in-time observational assessment of disease burden at a single point in time, with or without a longitudinal aspect, that is not a cohort study. |

The definition of a prospective cross-sectional prevalence study has changed over time. Originally, the word ‘prospective’ implied a future study event. Today, semantic broadening—due perhaps to the increasing presence of digital clinical data and, concomitantly, retrospective cross-sectional designs—has led to many non-retrospective cross-sectional prevalence studies being designated as “prospective”, even if they have no future component. As a result, the current most frequent, normative definition of a prospective cross-sectional prevalence study is simply a non-retrospective cross-sectional prevalence study that is clearly not longitudinal. The eligible publications in this scoping review’s random sample included frequent mention of “prospective” designs—all of those studies measured participants only once and had no longitudinal component. Mostly, this review uses the term prospective cross-sectional study both to ensure readers will not imagine the findings apply to retrospective studies, and to match its use in the publications included, as a normative descriptor of the traditional cross-sectional prevalence study.

### Benefits of prospective cross-sectional prevalence studies

Prospective cross-sectional prevalence studies are one of the most common study types performed in the world of global health. These studies have a number of benefits. They can help policymakers understand the investment needed to achieve improved health for specific diseases. Studies can help target investment toward the right populations and places. Further, prospective cross-sectional studies find and present evidence about subpopulations and variables that are associated with the most need, and may be associated with causal factors. In the absence of well-funded national surveillance or epidemiological programs, independent prospective cross-sectional prevalence studies provide evidence to help understand current states and future trends. Last but not least, these studies can increase the likelihood that new research and randomized controlled trials (RCT) are informative.

Human health research includes a variety of study types. The gold standard for evidence generation is today’s randomized clinical trial (Andre). That study type—regardless of trial design complexity—requires a recent and accurate measure of disease to estimate or optimize trial sites, duration, or costs. A prevalence estimate that is either sourced from too far away geographically, too old to be accurate, or measuring a disease definition or target population that is too different from the desired trial population, is likely to misrepresent the correct prevalence. As a result, recent, local cross-sectional prevalence studies represent a source of evidence critical to RCTs. This data influences biostatistical work on event rates, stratification, statistical simulation design and sample size efforts, as well as study design work on feasibility and trial site selection. If, for example, site prevalence is lower than estimates, the trial may never finish—a type of trial uninformativeness (Zarin).

### Rationale

This scoping review is targeted towards the “diseases of the poorest billion” (Coates). Diseases and conditions of the poorest billion map to a segment of the International Health Metrics and Evaluation (IHME)’s Global Burden of Diseases, Injuries and Risk Factors Study (GBD) (Vos). This segment is exclusively defined as categories of communicable, maternal, neonatal, and nutrition (CMNN). In a prospective cross-sectional prevalence study of the “diseases of the poorest billion” (Coates) in low-resource settings, the data or sample collection is “shoulder to shoulder” with the human participant that might have a disease. This sample data collection in studies varies widely depending on multiple factors. At times, individual households are randomized, and front-line health workers go to the door of the household to solicit informed consent and measurement. Alternately, a study may look for participants exclusively in institutions such as hospitals or schools. Finally, sometimes study workers will engage participants at workplaces or semi-public settings. Variables of interest to this review included data collection methods, diseases and conditions, presence of funding, types of ethics review, countries and sample sizes. The variety of data captured across study publications and lack of multiple datasets to power *a priori* questions drove the conclusion that introduction of a novel topical dataset lent itself to a scoping review approach.

The rationale of the current scoping review is to create a dataset to help understand the current state of human prospective cross-sectional prevalence studies in sub-Saharan Africa (sSA) focused on broad categories of diseases and conditions. No similar studies have been published. This scoping review’s topic of interest is not singular in any dimension other than a narrow focus on study design. This scoping review offers two views into the dataset. First, we provide a descriptive analysis to characterize a random sample of the larger dataset of studies identified for inclusion. Second, we present a collection of exploratory demonstration questions determined during full-text screening and data extraction, to illustrate the potential utility of interrogating such a dataset.

Noted biostatistician Miguel Hernan wrote about times it is acceptable or encouraged to ignore ‘power’ when introducing a novel dataset (Hernan). “When a causal question is important, it is preferable to have multiple studies with imprecise estimates than having no studies at all (Hernan).” In the vein of being the first of what Hernan would say is enough datasets to enable answering a question in a powered or meaningful way, the current scoping review is “provisional evidence” and will wait for other datasets to follow before questions will be appropriately powered (Hernan). As a result, this scoping review was not designed to answer *a priori* questions. This scoping review was designed to generate a dataset, to characterize a random sample of the dataset, and provide statistical insights to a basket of hypothetical demonstration questions covering an array of topics that might be of common interest.

#### Context in general

The search was limited to include four years’ worth (mid 2019-mid 2023) of prevalence studies conducted within sSA. This time period overlapped but did not completely encompass COVID-19. COVID-19 affected the clinical research landscape (London), so studies designed to measure COVID were excluded. Only studies that included a measure of disease prevalence in humans were included. Particular complexity arose when implementing the broad scope of including all diseases in the CMNN categories defined by IHME. The conditions and diseases in CMNN represented thousands of ICD-10 codes that authors needed to apply precisely in screening. These criteria together might be described as: if there was a one-time study with a true prevalence design, where humans were measured hands on to calculate traditional global health disease burden, happening anywhere in sSA in the last four years, that study publication is in this dataset.

#### Context examples

No scoping reviews exist for prospective cross-sectional prevalence studies implemented outside of national surveillance programs for multiple communicable disease categories, across Africa. Amongst the contextual examples identified, other scoping reviews pick a single disease, or a single target population, or a single influencing dimension (e.g., vector, intervention) and allow for breadth in other dimensions. One contextual example for this scoping review is for a single disease, with a focus on non-humans: “A scoping review on the prevalence of Shiga-toxigenic Escherichia coli in wild animal species” (Espinosa). Another focuses on communicable and non-communicable disease, and extends beyond sSA: “Chronic respiratory disease surveys in adults in low- and middle-income countries: A systematic scoping review of methodological approaches and outcomes (Hanafi).” A final example is a scoping review not about disease burden, but rather interventions: “Antimicrobial stewardship in South Africa: A scoping review of the published literature” (Chetty). None of these example studies would be eligible for inclusion.

### Objectives

The objectives of this scoping review include, for a single output dataset, to:

1. Create a dataset for prospective cross-sectional studies in diseases traditionally considered “global health” as defined according to a reproducible taxonomy limited to communicable, maternal, neonatal and nutritional, where these diseases and conditions describe the object of the prevalence measure, not the target population’s existing diseases or conditions
2. Create a dataset of studies that are prospective, cross-sectional prevalence type, where a human is part of the target population
3. Create a dataset where the intention is to have normative, limited in time, traditional studies to calculate current percentages of burden, and exclude retrospective studies and studies that are from national surveillance or epidemiological efforts
4. Create a dataset to find recent studies published in English that are open-access
5. Create a dataset for studies only taking place in sub-Saharan Africa (sSA)
6. Create a dataset that could be described or analyzed from a random sample rather than coding the entire dataset
7. Create a dataset, publish it in open access, and include accessible data
8. Describe a subset of the data extractable from study publications and how it can be used to answer questions on the topic concept.

## Methods

### About this review

The methodology for this scoping review was developed based on the 2020 PRISMA-SCR reporting guidelines (Tricco). This was supported by conceptual recommendations from the Joanna Briggs Institute Manual for Evidence Synthesis (Aromatis). A complete PRISMA-SCR 2020 checklist is included in the supplementary material or at the end of this paper (Tricco). As Tricco et al describe, “[Scoping reviews] may examine the extent (that is, size), range (variety), and nature (characteristics) of the evidence on a topic or question (Tricco).” In this scoping review, we will be examining characteristics of evidence on a topic, not a question. There was no *a priori* research question identified to drive this review. The topic driving the review was providing a dataset to power future analysis of the state of practice of cross-sectional prevalence studies in sSA in CMNN diseases. There was no protocol written for this scoping review, nor was this scoping review registered.

### Information Sources & Search

The literature search was conducted in a single database, PubMed. Due to the broad scope of the diseases included, the search was limited to publications that were indexed to the MeSH vocabulary for the relevant disease terms. “MeSH (Medical Subject Headings) is the National Library of Medicine controlled vocabulary thesaurus used for indexing articles for PubMed (MeSH).” An extensive mapping exercise was conducted to map each IHME CMNN disease term/category to a MeSH term in PubMed to be used in the search. The search was limited to publications including the word ‘prevalence’ in the title, and those published from June 1, 2019 to June 1, 2023. The date of search that produced the data was June 30, 2023, and that was the most recent date of any search. No language restrictions were applied in the search strategy. High volumes of appearances of the following terms within irrelevant publications identified in pilot searches led them to be filtered out using the search strategy: ‘COVID-19’, ‘retrospective’, ‘India’, ‘Pakistan’, and ‘cattle’. The complete search strategy is included in the supplementary materials. Because only one database was searched, no method for resolving or removing duplicates was needed; no duplicates were found.

### Study selection

#### Inclusion Criteria

Publications with all of the following criteria were included:

1. Studies conducted entirely within sSA: Studies must have all sites in sSA and no other site locations. African Union definitions of sSA countries were used. Studies with sites in India, Pakistan and Uzbekistan were incorrectly included; India and Pakistan studies were removed within study selection, and Uzbekistani studies were removed during screening.
2. Cross-sectional prevalence studies measuring the prevalence of a CMNN disease according to the categories defined by IHME GBD.
3. Studies measuring diseases and conditions in humans.

#### Exclusion Criteria

Publications with any of the following criteria were excluded:

1. An individual disease or condition whose prevalence is being measured where that individual disease’s ICD-10 code does not map to the ICD-10 code categories included in the IHME GBD’s CMNN segment (Diseases, WHO)
2. Studies designed to test an intervention or device (e.g., an innovative diagnostic or at-scale genomic testing)
3. Studies that were designed to test adherence to an intervention
4. Studies that were part of a single or ongoing national or subnational government surveillance, epidemiology or other structured health program
5. Studies with retrospective design (e.g., using secondary use electronic health record data and mining that data for historical prevalence)
6. Studies with study designs that have some longitudinal aspect, including designs that were longitudinal, cohort, or repeated measures designs
7. Studies with designs that were comparative in nature, where one cohort’s prevalence is compared with another cohort’s prevalence, specifically to inform analyses or action related to an intervention
8. Studies with study designs that were case reports or case-control studies
9. Studies with study designs that were systematic reviews or meta-analyses
10. Publications that were abstracts, posters, or protocols
11. Approaches where the exclusive prevalence target population (i.e., the target population is being measured for whether they have a disease or condition) is non-human (e.g., rodents, dog, cattle, a countertop or surface)

**Table 2.**
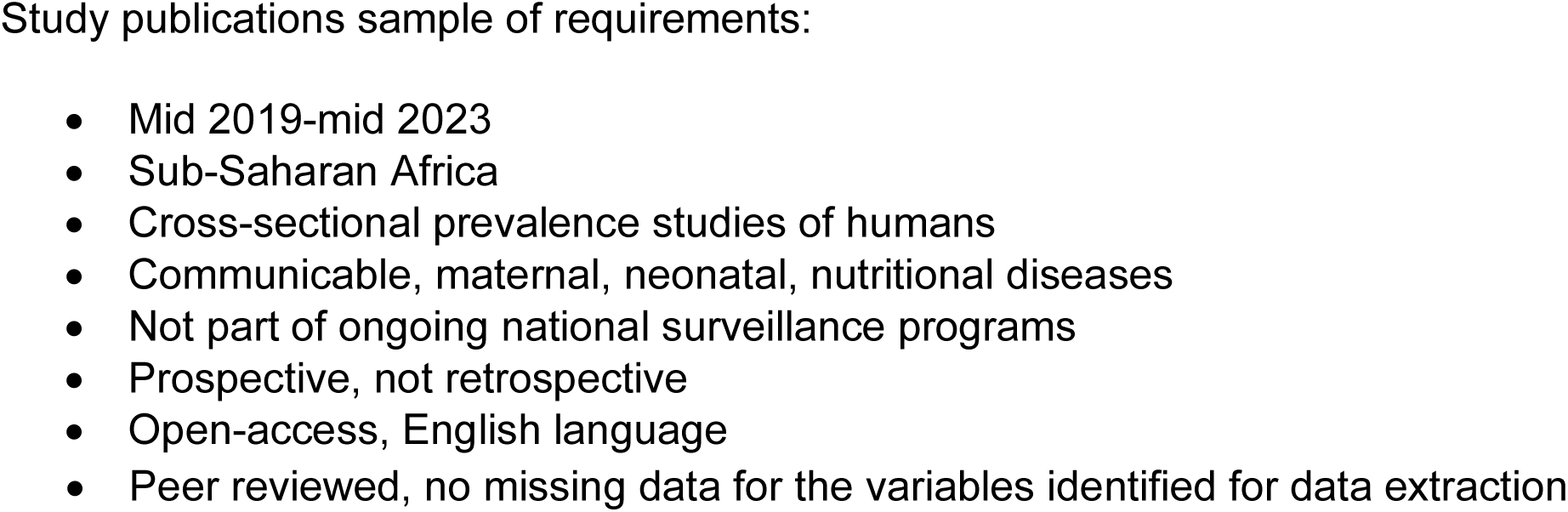
Snapshot of consolidated criteria for inclusion & eligibility.

### Screening

#### Selection of Sources of Evidence

Search results were imported into Rayyan.ai (Boston, Massachusetts, USA) for screening. Titles and abstracts were independently screened by two reviewers (BB and SD) to determine eligibility, according to the inclusion and exclusion criteria. A mapping tool for use with World Health Organization (WHO) International Classification of Diseases (ICD) codes (WHO) for CMNN and other diseases, assembled by the IHME, was utilized (Diseases). Disagreements were resolved by consensus. Rarely, full texts of publications were consulted to clarify challenging language in the abstracts. Comprehensive full-text screening was conducted on a random sample of studies eligible for inclusion based on abstract screening. The identification of a random sample for data extraction is described in detail below. Full-text screening on the random sample of studies was conducted by a single author (SD).

### Data extraction

#### Creating and coding a random sample

Without the time and resources to code all potentially eligible publications, this scoping review opted to code and analyze a random sample likely to be representative of those publications. This approach might be non-normative for a research question-based traditional scoping review, in the presence of prior evidence. However, in the context of an absence of other evidence, and a lack of driving research questions requiring formal power calculations, it was determined that this topic-based expository of novel observational data could achieve its goals without full data coding. This approach has been used before in a contemporary scoping review (Leung).

The minimum sample size of publications coded was set at 30 to optimize the sample’s likelihood to be representative of the larger dataset. Thirty was identified as likely to generate reasonable confidence that any inferences made from the random sample would be likely to generalize to the full sample.

The sample size of 30 was selected for its sampling properties, not for power levels for individual hypotheses. “If a sample consists of at least 30 independent observations and the data are not strongly skewed, then the distribution of the sample mean is well approximated by a normal model (Vu).” “When the sample size is 30 or more, we consider the sample size to be large, and by Central Limit Theorem [the sample mean] will be normal even if the sample does not come from a Normal Distribution. Thus, when the sample size is 30 or more, there is no need to check whether the sample comes from a Normal Distribution (Department of Statistics).”

To create the sample, publications were assigned into a random order from the overall dataset by the biostatistician (CM). These publications were serially screened by a single reviewer (SD). Publications were screened and coded in the randomly assigned order until the quantity of eligible publications reached 38 in total. Coding 38 eligible publications was required to provide a set of 30 publications that met a key component of a hypothetical demonstration question of interest.

From time to time, publications included information in their full-text versions that was not represented in their abstracts. As a result, full-text publications in the random sample were assessed to identify if they matched the inclusion/exclusion criteria from the screening phase. This assessment found three studies that would have been excluded in a prior phase were more data present in the abstracts. One of the three included a retrospective design, and two were routine parts of large national epidemiology programs. All were excluded. An additional publication was excluded as it was not available in open access. Four publications were excluded as each was missing data that precluded coding. In total, eight studies were excluded.

Upon review, if a single publication presented results from more than one appropriately designed study, only the most recent study was included in the analysis to avoid overweighting the results of a single paper and to maintain independence across studies. One instance of this occurred in the random sample.

When analyzing the data from the random sample, the distributions of several continuous variables of interest were found to be right-skewed, so all differences between groups were evaluated using Wilcoxon rank-sum tests. The use of this approach obviated the need to remove outlier data. Categorical data were analyzed using Fisher’s exact test. No formal power calculations were planned. Several questions of interest involved the dynamics of prevalence studies conducted in more general populations, so we sought to include a minimum of 30 publications that were not specifically restricted to pregnant women. A total of 38 publications needed to be coded to reach this threshold. The random sample includes 38 publications in total.

Outside of the random sample, full texts for all other publications were retrieved. During full text retrieval, publications not available in open access were excluded. Further, a publication with an English abstract but only French full text publication was excluded. No authors or investigators were contacted at any stage of this review.

Additional eligibility criteria applied during data extraction were limited to a) publications must be open access, so as many researchers as possible can leverage the entire dataset, b) publications must have an English language full-text publication in addition to any local language full-text publications, c) full-text publications must report key information consistently throughout the publication (e.g., if the paper in the abstract reports data collection happening April to June, and in the methods section reports data collection happening December to February, then it is impossible to know which is correct), and d) full-text publications must include all key variables identified for data extraction (e.g., if it does not describe or announce the method of data collection or when the journal received the article, it cannot contribute to a larger dataset that is complete). The one exception for ‘d’ is that there is a convention in some journals—especially journals from Africa—that when there is no funding for a study, the journal will not require authors to document that fact. Instead, the lack of information about funding clearly means no funding was utilized for the study. Absence of funding information was not considered missing data.

#### Coding the random sample

After reviewing all potentially eligible publications for presence of open access full texts, and identifying exclusions during full-text screening of the random sample, the result was 363 open access peer-reviewed English publications describing cross-sectional prevalence studies as defined herein. Assessing the random sample full text publications, a set of 16 raw data items (Table 3) and 9 aggregate data items (Table 4) were selected. Aggregate data items were ones on which calculations, summarizations, or classifications were applied to improve the quality of reporting, visualizations, and further analysis.

The fully coded dataset is available in the supplementary material.

#### Data Items

**Table 3:**
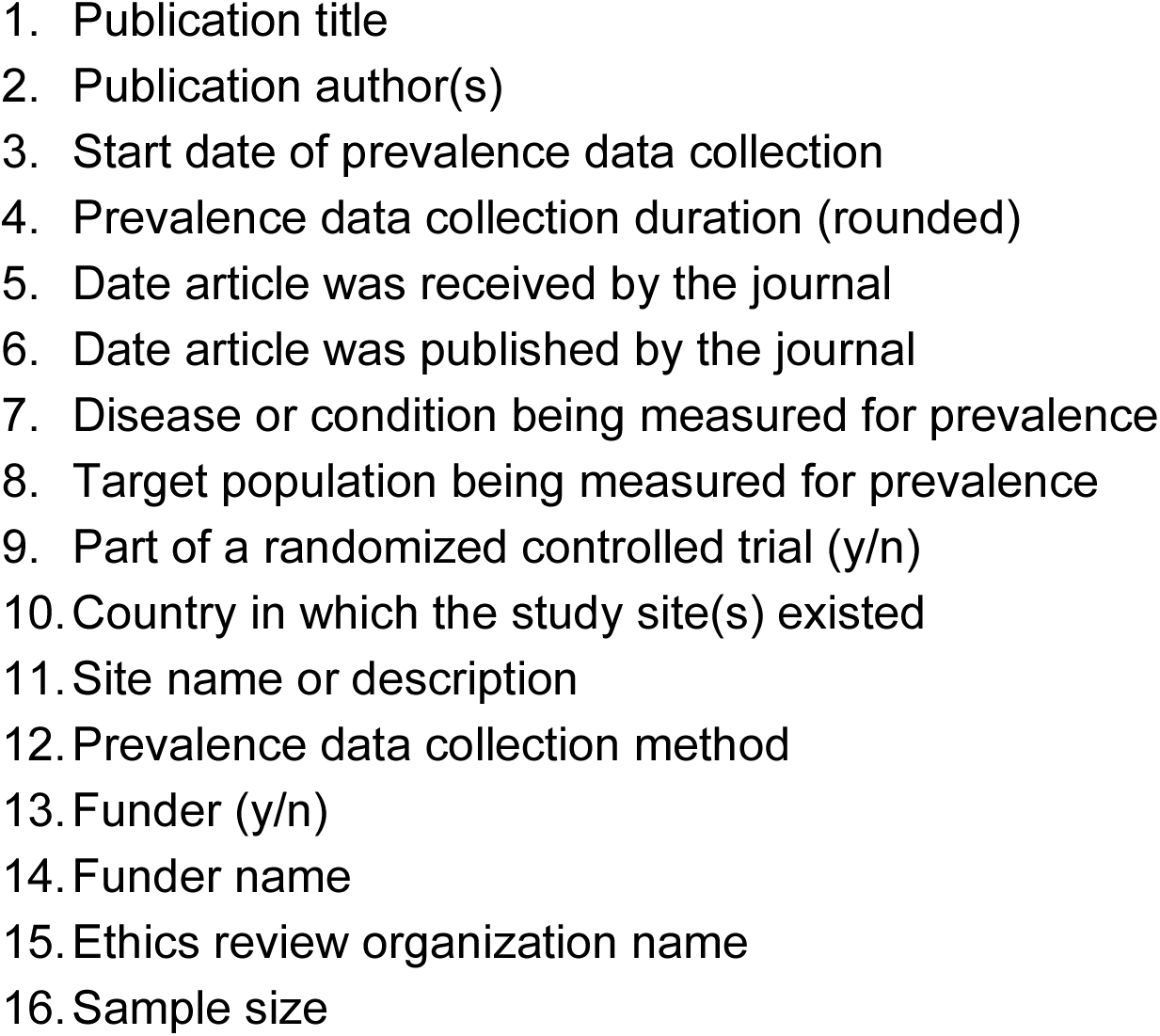
Original raw data variables coded from a random sample of the dataset.

**Table 4:**
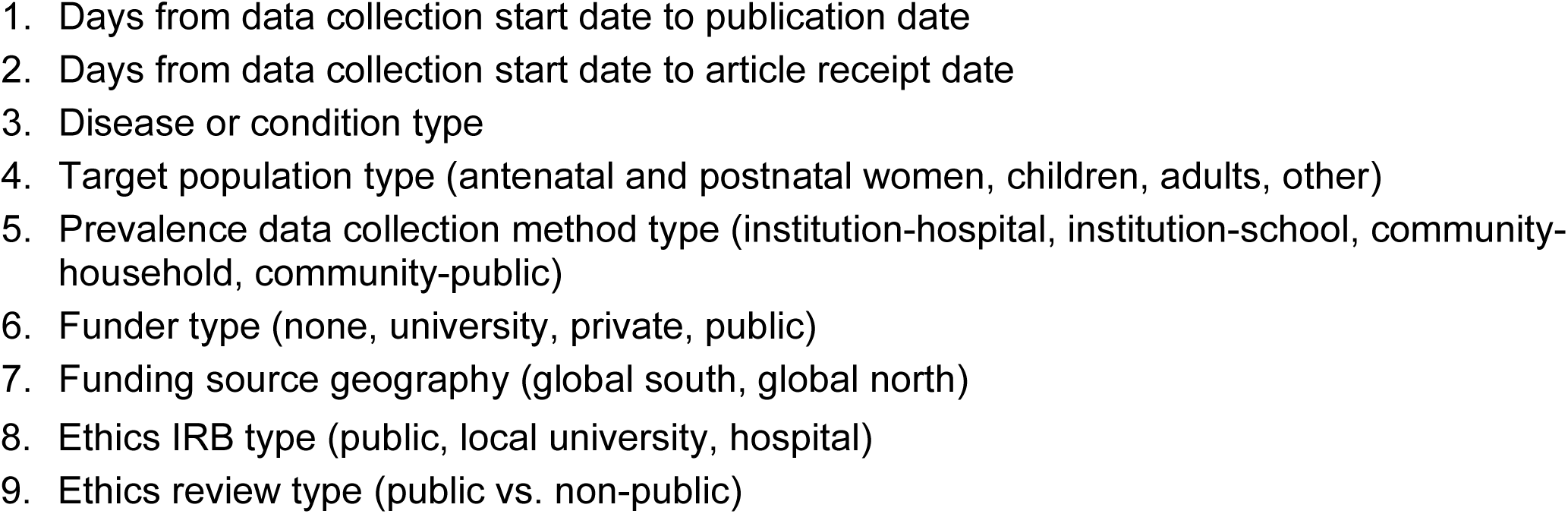
Variables using calculations or summarizations of original data, with examples.

#### Characterizing the raw, calculated, and summarized data

The random subset of the dataset that was coded included information on 25 variables (Tables 3, 4). Of the 25 variables, 8 variables were selected as the most meaningful to characterize (Table 5). Charts regarding and describing the 8 variables are described in the Results section.

**Table 5:**
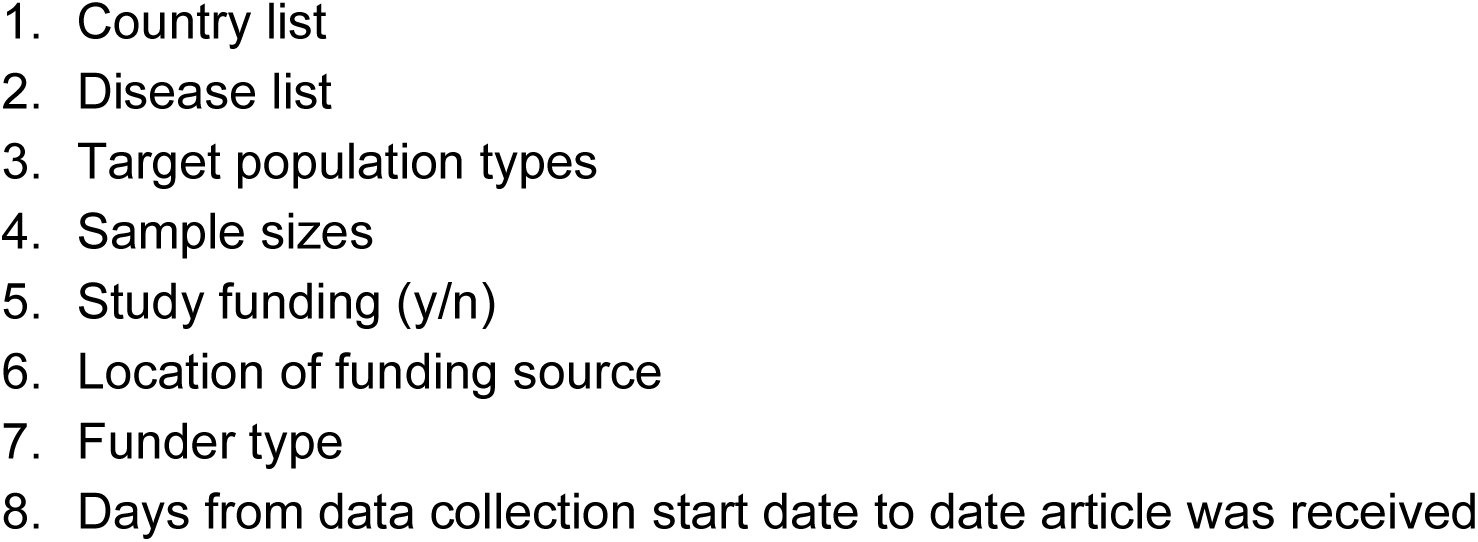
Dataset variables most meaningful to characterize.

### Data analysis

#### Characterizing a sample of demonstration questions statistically

A random selection of potentially meaningful questions was posited after screening. SD and CM created 11 questions and their null hypotheses. The questions were designed to aid in one of the objectives of this review: describe a subset of the data extractable from study publications and demonstrate the potential utility of that data to answer questions on the topic concept. While the rationale for these questions was not to be ‘well powered’ (Hernan), this review nevertheless provides p-values to characterize the strengths of associations in the random sample and to offer suggestions for future statistical evaluation were the entire dataset coded and analyzed. Caution should be exercised when interpreting the results since this scoping review may not have had a large enough sample size to reliably detect the presence of certain effects or relationships. CM performed the analysis. Demonstration questions and their null hypotheses are listed in Table 6.

**Table 6.**
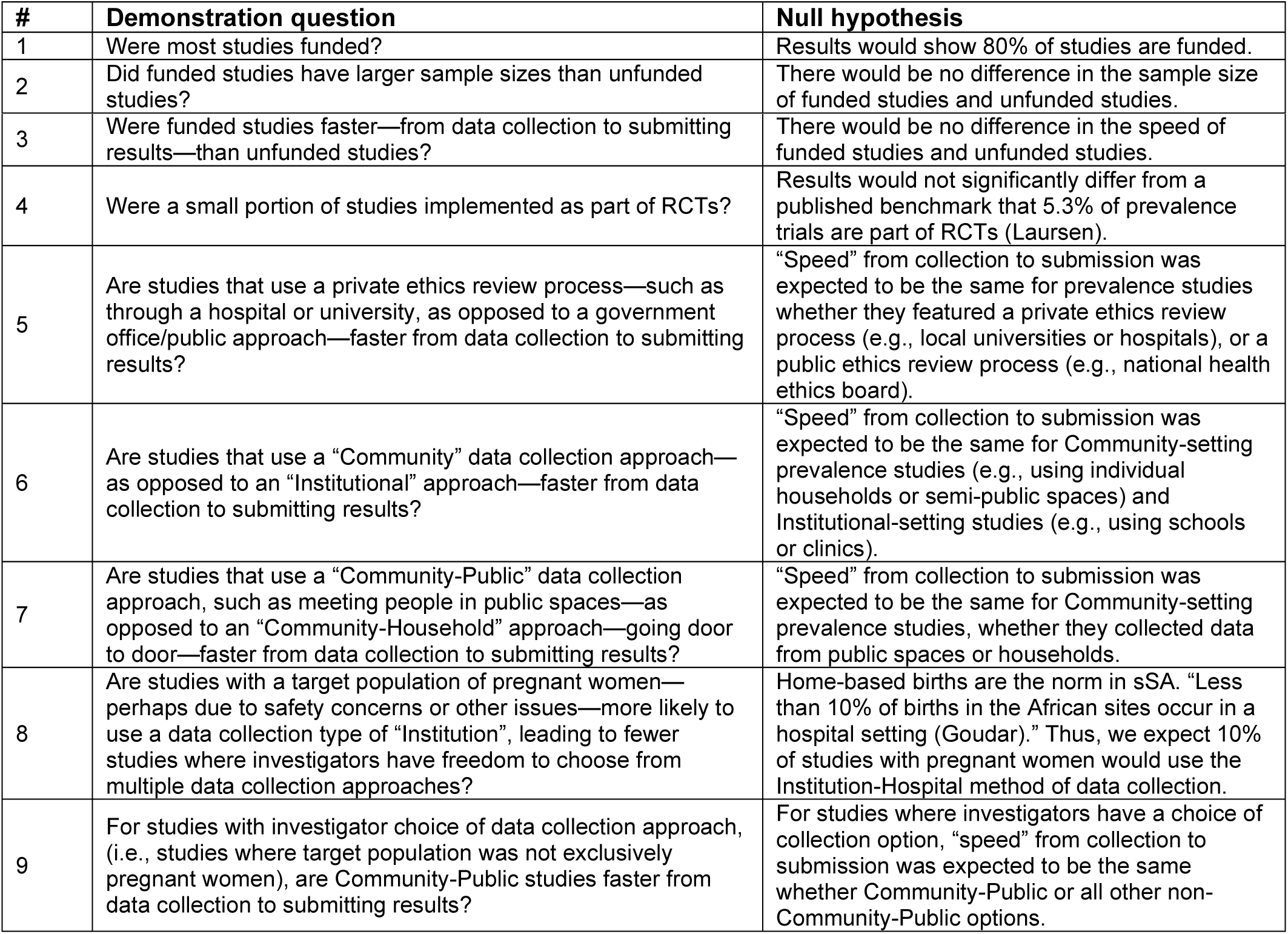
Hypothetical demonstration questions applied to the dataset.

| # | Demonstration question | Null hypothesis |
| --- | --- | --- |
| 1 | Were most studies funded? | Results would show 80% of studies are funded. |
| 2 | Did funded studies have larger sample sizes than unfunded studies? | There would be no difference in the sample size of funded studies and unfunded studies. |
| 3 | Were funded studies faster—from data collection to submitting results—than unfunded studies? | There would be no difference in the speed of funded studies and unfunded studies. |
| 4 | Were a small portion of studies implemented as part of RCTs? | Results would not significantly differ from a published benchmark that 5.3% of prevalence trials are part of RCTs (Laursen). |
| 5 | Are studies that use a private ethics review process—such as through a hospital or university, as opposed to a government office/public approach—faster from data collection to submitting results? | “Speed” from collection to submission was expected to be the same for prevalence studies whether they featured a private ethics review process (e.g., local universities or hospitals), or a public ethics review process (e.g., national health ethics board). |
| 6 | Are studies that use a “Community” data collection approach—as opposed to an “Institutional” approach—faster from data collection to submitting results? | “Speed” from collection to submission was expected to be the same for Community-setting prevalence studies (e.g., using individual households or semi-public spaces) and Institutional-setting studies (e.g., using schools or clinics). |
| 7 | Are studies that use a “Community-Public” data collection approach, such as meeting people in public spaces—as opposed to an “Community-Household” approach—going door to door—faster from data collection to submitting results? | “Speed” from collection to submission was expected to be the same for Community-setting prevalence studies, whether they collected data from public spaces or households. |
| 8 | Are studies with a target population of pregnant women—perhaps due to safety concerns or other issues—more likely to use a data collection type of “Institution”, leading to fewer studies where investigators have freedom to choose from multiple data collection approaches? | Home-based births are the norm in sSA. “Less than 10% of births in the African sites occur in a hospital setting (Goudar).” Thus, we expect 10% of studies with pregnant women would use the Institution-Hospital method of data collection. |
| 9 | For studies with investigator choice of data collection approach, (i.e., studies where target population was not exclusively pregnant women), are Community-Public studies faster from data collection to submitting results? | For studies where investigators have a choice of collection option, “speed” from collection to submission was expected to be the same whether Community-Public or all other non-Community-Public options. |

No critical appraisal of the data source was performed.

### Synthesis of data results

The methods of handling and summarizing the data that was extracted include the following. First, after eligibility and coding was performed for the granular data items in the random sample of 38 publications, aggregate categories or ‘types’ were created for possible data items. These type values were based on easy-to-understand, qualitative, bottom-up approaches. Type columns were populated (Microsoft Excel, Redmond, Washington, USA). Next, data was provided for analysis related to demonstration questions. Those analyses were performed in R version 4.2.3 (RStudio, Vienna, Austria). Finally, the descriptive reports were created in Microsoft Power BI (Redmond, Washington, USA).

Methodological quality assessments and within-study risk of bias were not assessed due to limited time and resources, and the novel nature of the data. Per the PRISMA-SCR checklist, risk of bias across studies and additional analyses are not applicable for scoping reviews.

## Results

### Selection of sources of evidence

See Figure 1, the PRISMA-SCR flow diagram to identify sources of evidence assessed during phases of Search, Screening, and Eligibility.

**Figure 1.**
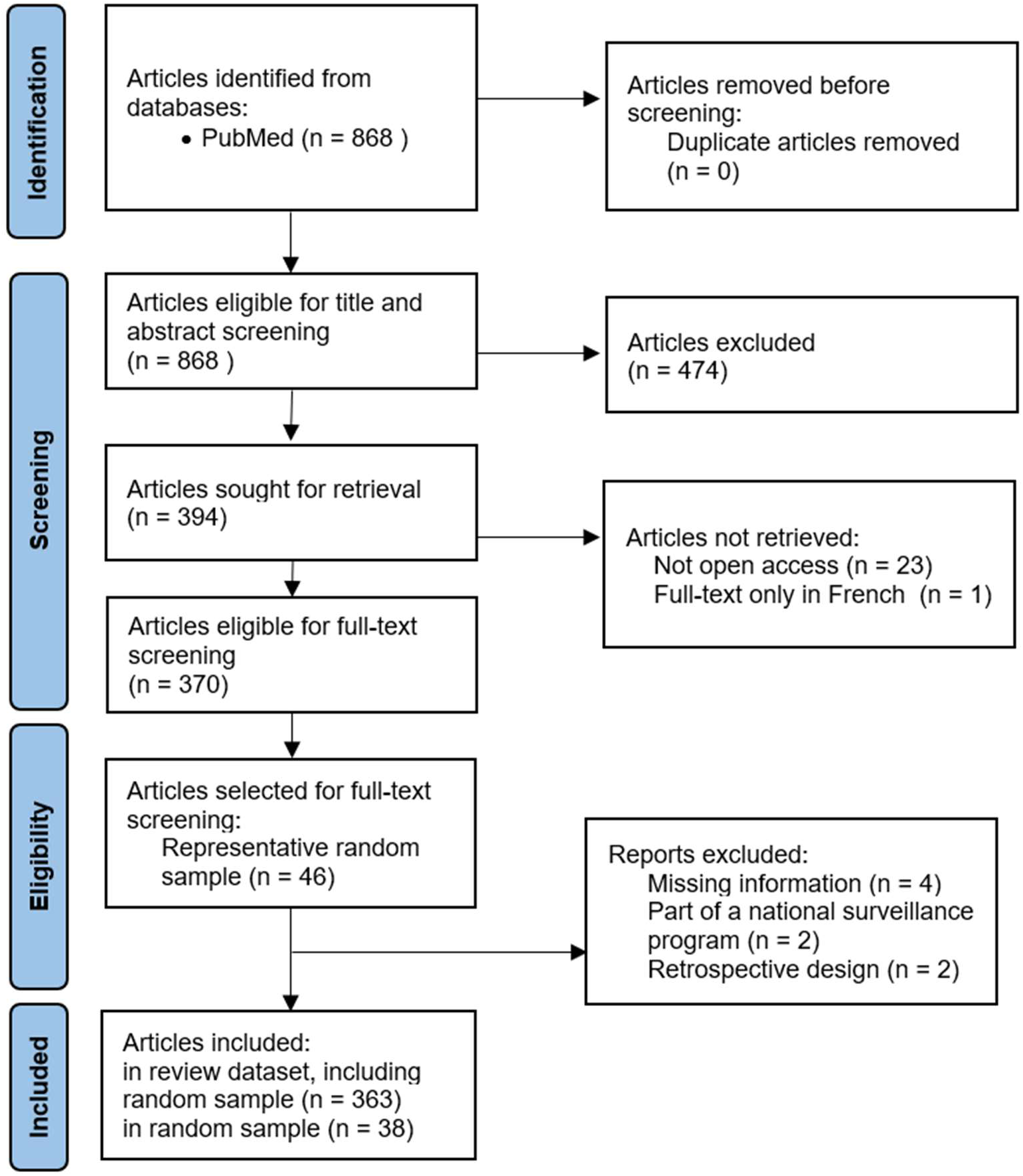
PRISMA flow diagram, adapted to accommodate random sample-.

### Characteristics of sources of evidence

Descriptive statistics of the screened publications is in Table 7. Lack of full dataset coding precludes complete descriptive statistics on all screened and eligible publications.

**Table 7.** Descriptive statistics.

| **Characteristic of publication(s) in dataset prior to screening** | **Count of publications** |
| --- | --- |
| **Total** |  |
| Non-excluded publications | 394 |
| **Year of publication** |  |
| 2019 (partial) | 64 |
| 2020 | 107 |
| 2021 | 98 |
| 2022 | 99 |
| 2023 (partial) | 26 |
| **Journal (minimum 15 publications)** |  |
| PLOS One | 83 |
| BMC Infectious Diseases | 23 |
| Pan African Medical Journal | 19 |
| Malaria | 18 |
| PLOS Neglected Tropical Diseases | 18 |
| BMC Pregnancy & Childbirth | 16 |
| BMC Public Health | 15 |
| **Condition keywords (minimum 10)** |  |
| Malaria | 56 |
| HIV | 37 |
| Parasitical | 31 |
| Intestinal | 27 |
| Hepatitis B | 18 |
| Anemia | 18 |
| Stunting | 14 |
| TB | 12 |
| *Schistosoma mansoni* | 12 |

Citations for all screened studies are in the supplementary materials.^S4:1-363^ No critical appraisal of included sources of evidence was completed. The first 38 studies in the supplementary references were the entirety of the random sample.^S4:1-38^

### Synthesis of results

Charts were created to describe the granular data and summarizations in the random sample, per the objectives. These are presented here as Tables 8-13. Tabular results from the hypothetical demonstration questions evaluated in the random sample are shown in Table 14.

Table 8 shows the countries in which the cross sectional prevalence studies occurred. All but one sample study had sites exclusively within one country. Only countries with two or more studies in the sample appear in Table 8.

**Table 8.** SSA countries with 2+ studies in random sample.

List of countries
| Countries | Studies |
| --- | --- |
| Ethiopia | 17 |
| Cameroon | 3 |
| Uganda | 3 |
| Mozambique | 2 |
| Somalia | 2 |
| Sudan | 2 |
| Tanzania | 2 |

Only countries with two or more publications in the sample are shown.

**Table 9.**
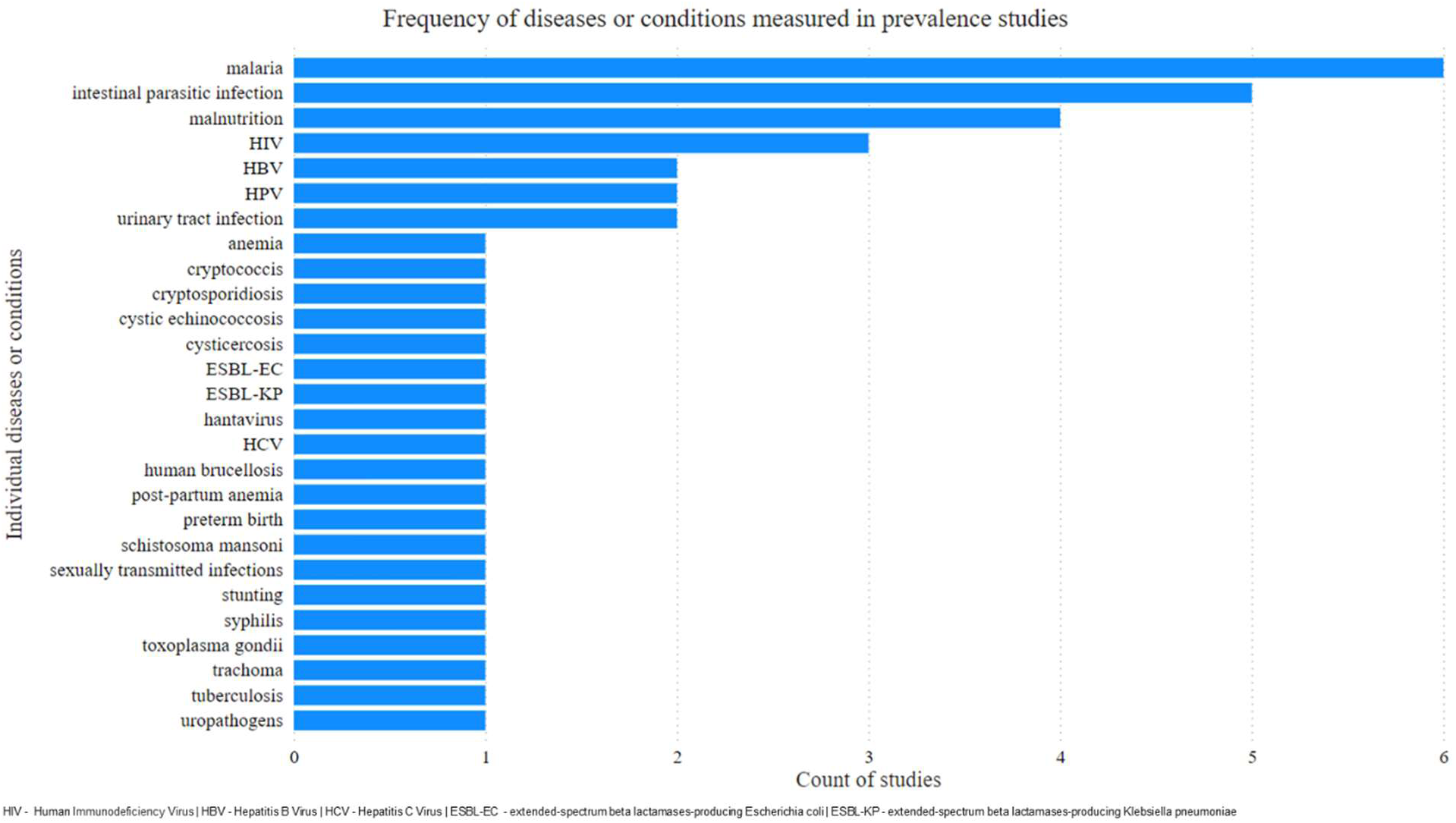
Individual diseases and conditions for which prevalence was measured.

Table 10 shows summary types of target populations. These were the groups that were measured for the presence or absence of a disease or condition. Age ranges varied. Some bespoke and specialized target populations appeared. These are included in the ‘Other’ category.

**Table 10.**
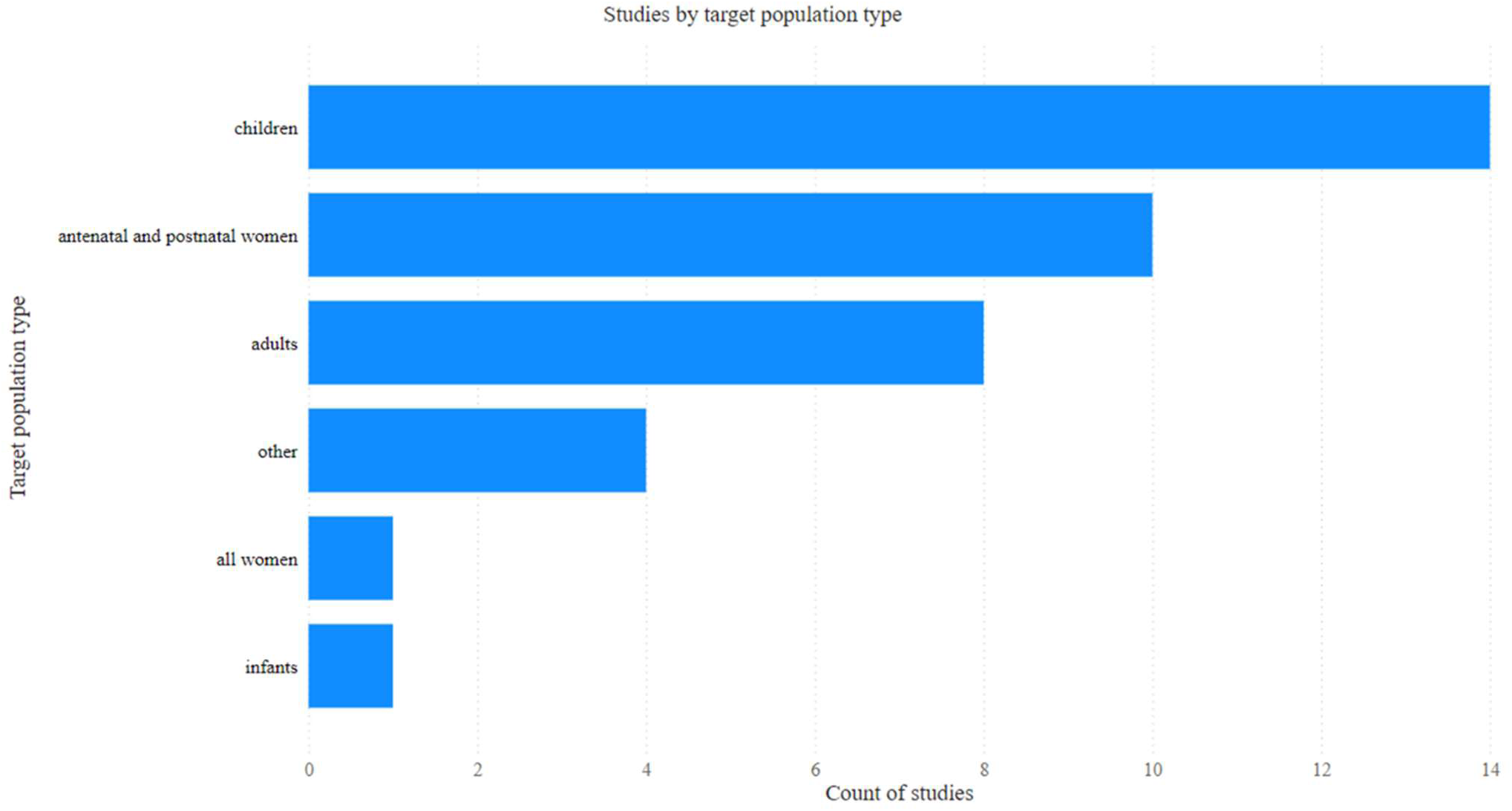
Target population types in the random sample.

Table 11 shows the sample size of the target populations in the sample studies. A large cluster of studies used sample sizes in the low hundreds. There were six studies with sample sizes in the thousands.

**Table 11.**
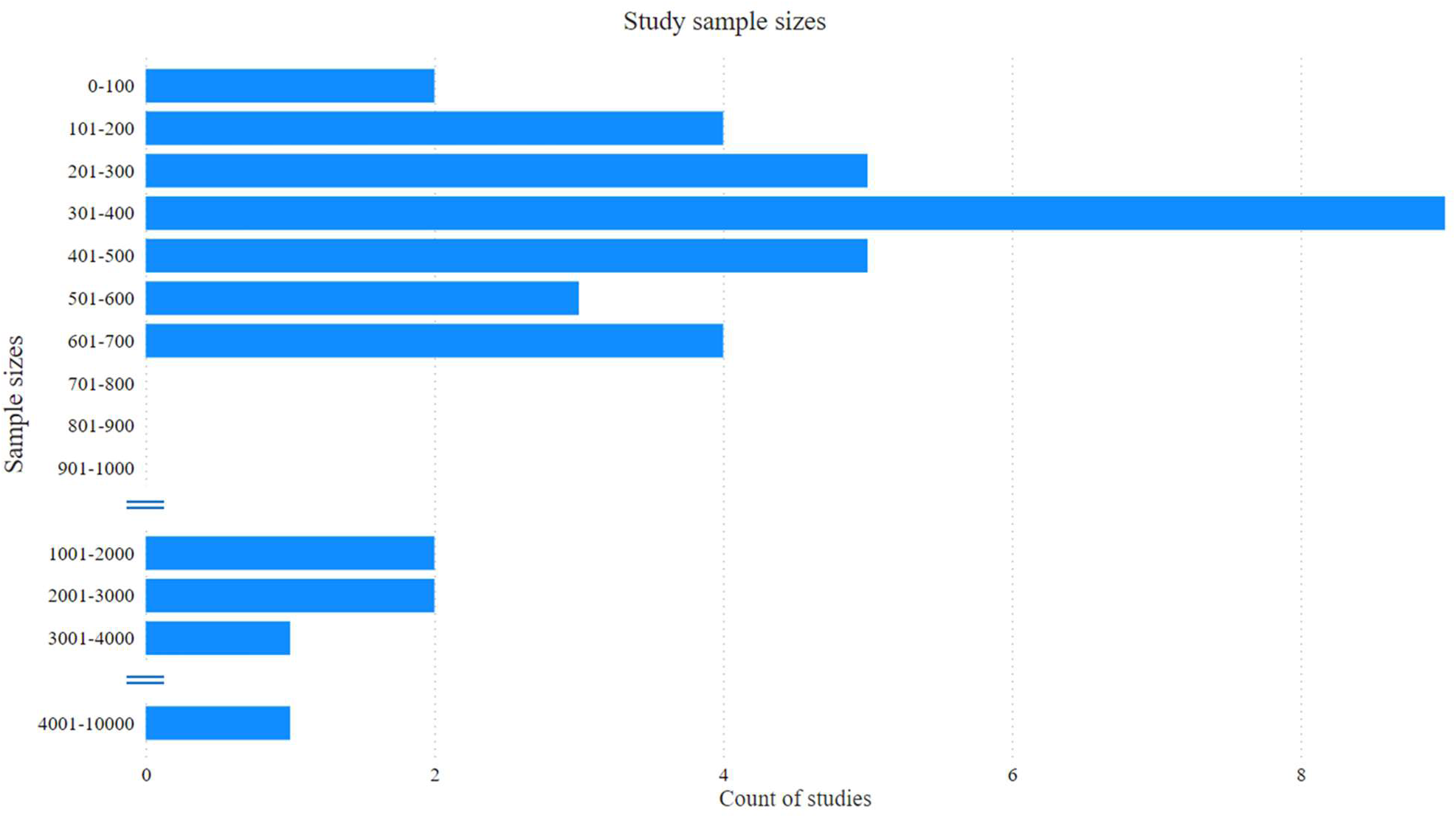
Target population sample size in the random sample.

Many studies were not funded. Of those funded, studies were about as likely to receive it from the Global South—in many cases local sources—as from the Global North. Table 12 shows three views into study funding.

**Table 12.**
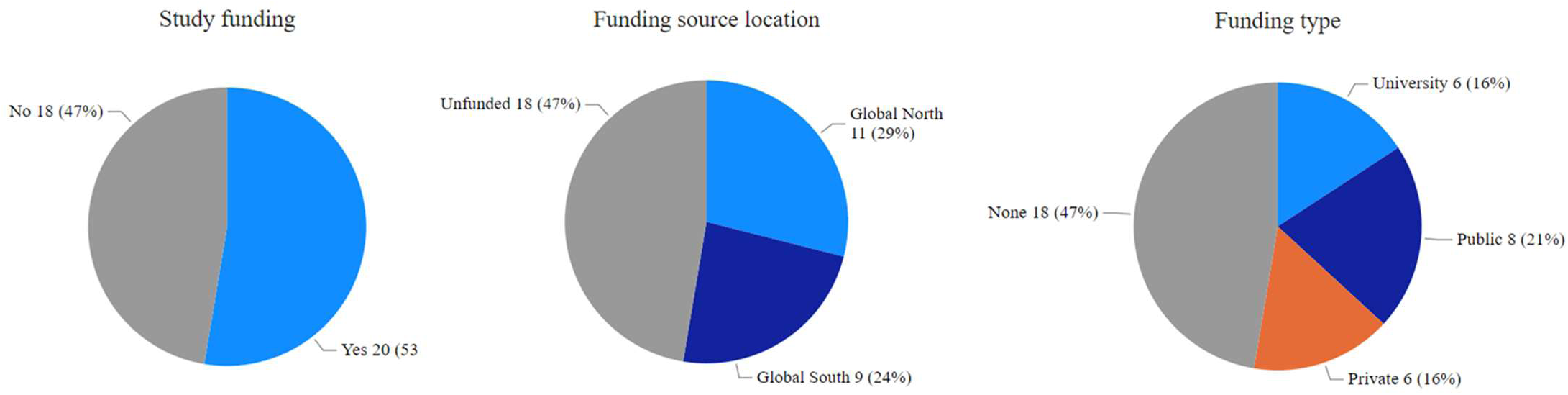
Funding and funding sources for studies in the random sample.

Table 13 shows the duration of a portion of the study span for studies in the random sample. The portion of the study span measured extends from the start date of their data collection with human participants to the date their article was received by the publishing journal. There is no way— outside of interviewing investigators—to create true ‘study start’ date; these studies are rarely entered in a trial registry. At the end of the duration, to neutralize across-journal variations in publication execution time, this review used the date the article was submitted by investigators to the journal. This study segment—from data collection start to article submission—should represent at least a majority of the study span. Table 13 also cross-tabulates these durations with the type of ethics institutional review board (IRB) that the study utilized. At times, these were central, national, regional or subregional government offices (public type) or local hospitals or universities (private).

**Table 13.**
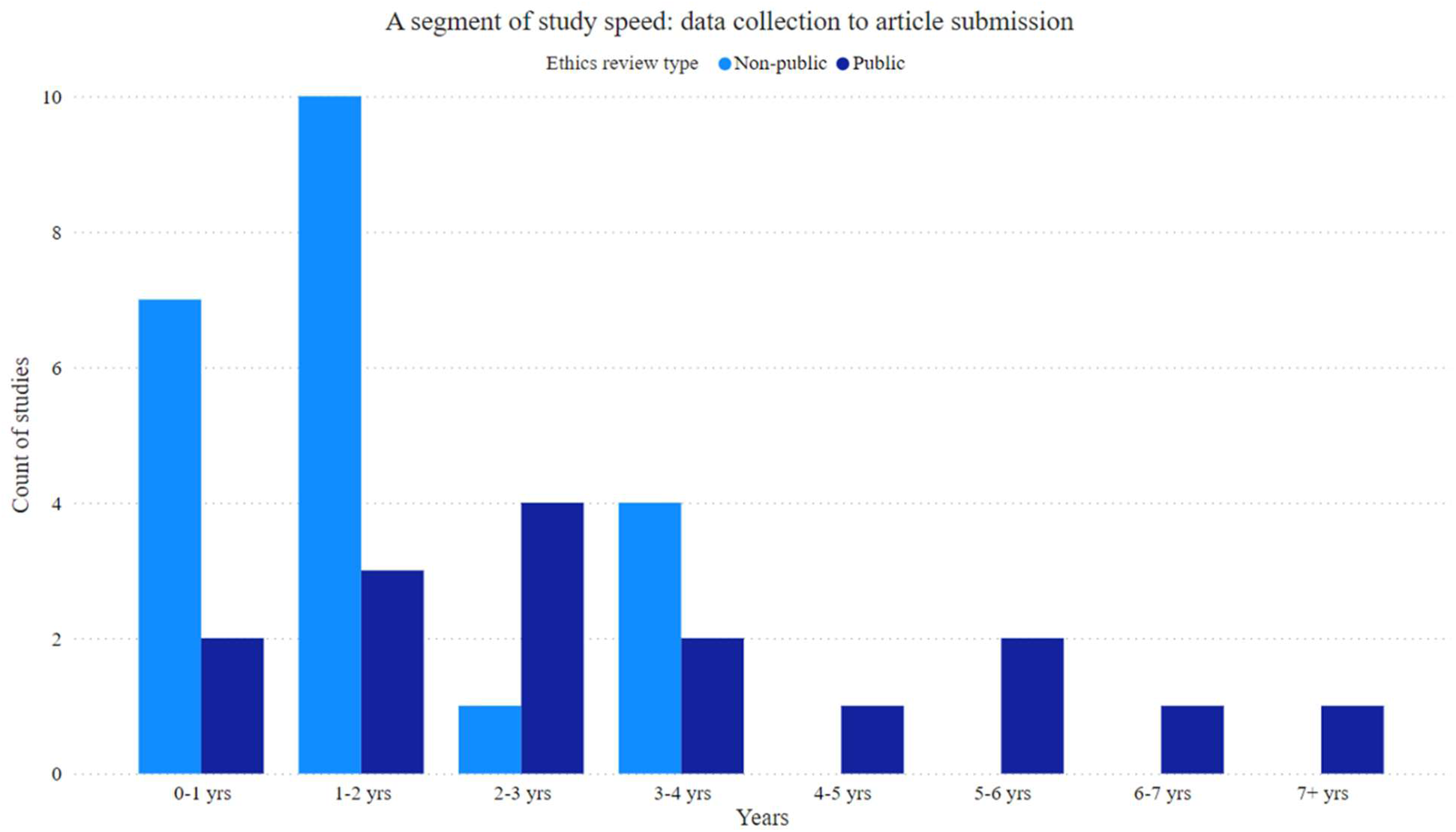
Duration from data collection to article submission by ethics review type.

### Hypothetical Demonstration Question Results

The demonstration questions in Table 6 are listed below, with results and p-values from hypothesis tests performed. Please note the scoping review was not powered to evaluate these demonstration questions, so hypothesis tests may be underpowered. These are exploratory analyses that were not specified *a priori*, but can point to potential future questions to be examined in a more rigorous way.

**Table 14.**
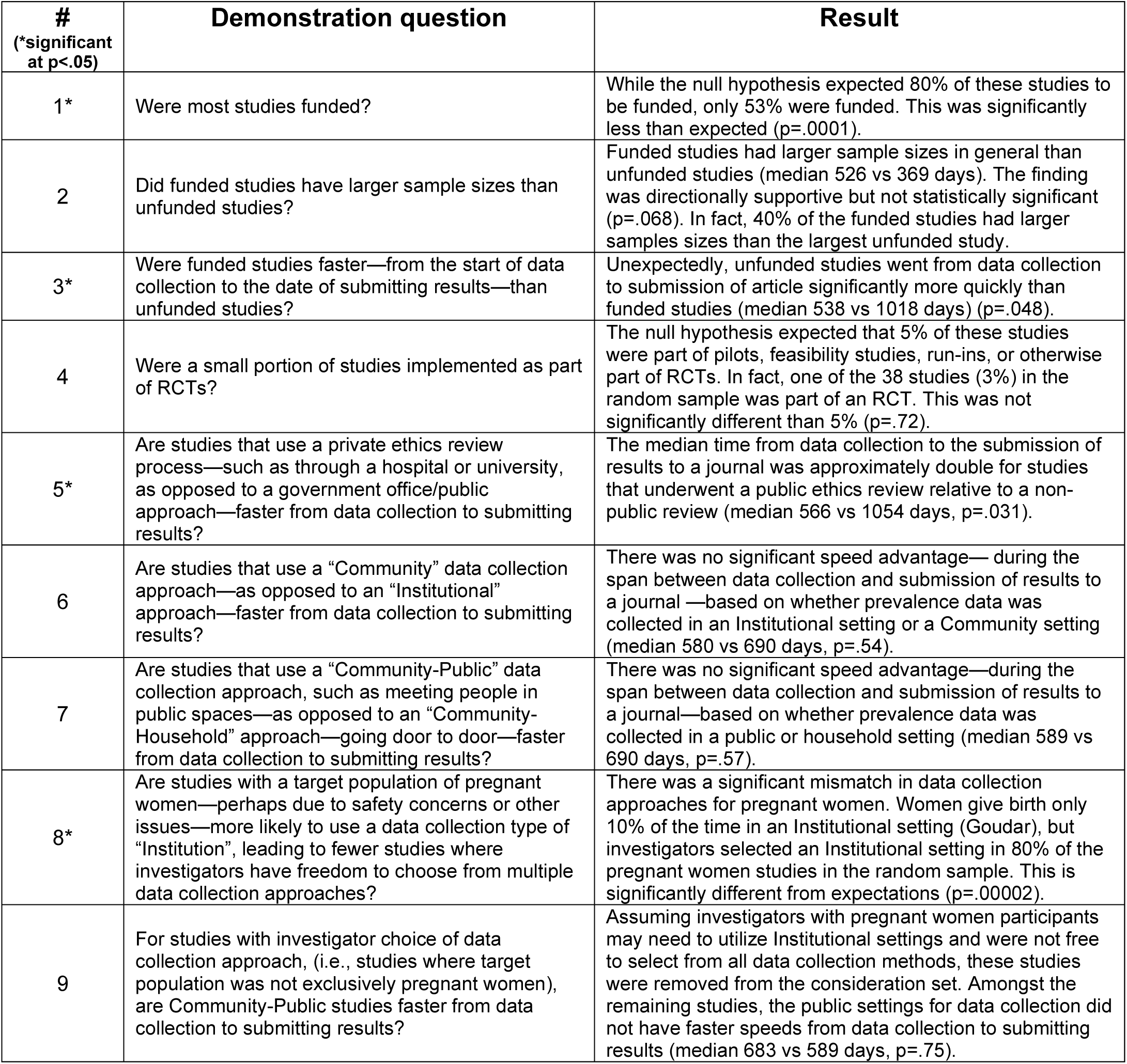
Hypothetical demonstration questions and resulting p values.

| #<br>(*significant<br>at $p < .05$) | Demonstration question | Result |
| --- | --- | --- |
| 1* | Were most studies funded? | While the null hypothesis expected 80% of these studies to be funded, only 53% were funded. This was significantly less than expected ($p = .0001$). |
| 2 | Did funded studies have larger sample sizes than unfunded studies? | Funded studies had larger sample sizes in general than unfunded studies (median 526 vs 369 days). The finding was directionally supportive but not statistically significant ($p = .068$). In fact, 40% of the funded studies had larger sample sizes than the largest unfunded study. |
| 3* | Were funded studies faster—from the start of data collection to the date of submitting results—than unfunded studies? | Unexpectedly, unfunded studies went from data collection to submission of article significantly more quickly than funded studies (median 538 vs 1018 days) ($p = .048$). |
| 4 | Were a small portion of studies implemented as part of RCTs? | The null hypothesis expected that 5% of these studies were part of pilots, feasibility studies, run-ins, or otherwise part of RCTs. In fact, one of the 38 studies (3%) in the random sample was part of an RCT. This was not significantly different than 5% ($p = .72$). |
| 5* | Are studies that use a private ethics review process—such as through a hospital or university, as opposed to a government office/public approach—faster from data collection to submitting results? | The median time from data collection to the submission of results to a journal was approximately double for studies that underwent a public ethics review relative to a non-public review (median 566 vs 1054 days, $p = .031$). |
| 6 | Are studies that use a “Community” data collection approach—as opposed to an “Institutional” approach—faster from data collection to submitting results? | There was no significant speed advantage—during the span between data collection and submission of results to a journal—based on whether prevalence data was collected in an Institutional setting or a Community setting (median 580 vs 690 days, $p = .54$). |
| 7 | Are studies that use a “Community-Public” data collection approach, such as meeting people in public spaces—as opposed to an “Community-Household” approach—going door to door—faster from data collection to submitting results? | There was no significant speed advantage—during the span between data collection and submission of results to a journal—based on whether prevalence data was collected in a public or household setting (median 589 vs 690 days, $p = .57$). |
| 8* | Are studies with a target population of pregnant women—perhaps due to safety concerns or other issues—more likely to use a data collection type of “Institution”, leading to fewer studies where investigators have freedom to choose from multiple data collection approaches? | There was a significant mismatch in data collection approaches for pregnant women. Women give birth only 10% of the time in an Institutional setting (Goudar), but investigators selected an Institutional setting in 80% of the pregnant women studies in the random sample. This is significantly different from expectations ($p = .00002$). |
| 9 | For studies with investigator choice of data collection approach, (i.e., studies where target population was not exclusively pregnant women), are Community-Public studies faster from data collection to submitting results? | Assuming investigators with pregnant women participants may need to utilize Institutional settings and were not free to select from all data collection methods, these studies were removed from the consideration set. Amongst the remaining studies, the public settings for data collection did not have faster speeds from data collection to submitting results (median 683 vs 589 days, $p = .75$). |

## Discussion

A total of 363 publications of select cross sectional prevalence studies in sSA were identified in this scoping review as eligible for inclusion in a novel dataset. These prevalence studies covered a wide variety of individual diseases and conditions, across at least 17 sSA countries. Studies measured human participants for disease over days, months, or years. Their ethics reviews took place in a variety of sSA venues, as well as in Europe and America. Sample sizes ranged from dozens to thousands. In our random sample of the dataset, study sites ranged from hospitals, clinics, blood banks, dialysis centers, schools, university cafeterias, households, and refugee camps.

The portion of these studies documented as being part of or in service to a future large clinical trial is marginal. It could be that RCT sponsors are funding cross-sectional prevalence studies that are retrospective or otherwise excluded from this review. Alternately, these sponsors may be funding studies and requesting no peer-review publication. It is more likely that RCTs are leveraging less local, less recent prevalence estimates that have been published organically and independently. A bibliometric study of where these prevalence publications are cited—either in published or registered study protocols, trial registries, or RCT publications—could identify downstream value.

A large portion of the studies were self-funded by principal investigators. These scientists and clinicians shouldered the burden of moving science forward in their regions and beyond. The Global North is a minority funder of these trials. There may be a ‘story’ here. The story includes a league of heroes who take up the prevalence questions and self-fund the answer, with symbolic but minimal support from the institutions around them. Use of their study results for positive health actions owe a debt to the PI’s key role as funder.

## Limitations

A number of limitations could be active in this scoping review:

1. While scoping reviews to map the evidence on a topic are somewhat well known (Tricco), taking a random sample of eligible studies for data extraction and analysis is a less common approach. Some authors, investigators, epidemiologists, drugmakers, students, and policymakers may be unfamiliar with this type of scoping review. This scoping review is a review of a topic, not a question (Tricco). Much of the value in this review lies in providing a first dataset toward a more powered ensemble (Hernan). More scoping reviews focused on topics and datasets, rather than research questions, if published, will broaden the perspective of readers and offer secondary research opportunities.
2. The search was limited to one database, and the search strategy for disease terms was limited to the MeSH vocabulary, meaning that some relevant publications would not have been found.
3. The IHME CMNN disease categories include multiple recursive hierarchies and are defined according to a complex and large map of ICD-10 codes. Using this system for disease definition opens the possibility of rating errors.
4. This review used the presence of the word ‘prevalence’ in the title of the publications as an inclusionary search term. While advance investigation showed this was rare, some studies could be appropriate prevalence studies, not have the word ‘prevalence’ in the title, and be missed.
5. This review covers four years of publication dates. That span includes a large proportion of COVID-19 years. COVID-19 changed or affected all aspects of the research ecosystem. As such, COVID-19 could have affected the research and publication processes for the studies in this review’s dataset.
6. While no research questions were specified *a priori*, coding the entire dataset would offer more precise descriptions of the dataset than using a random sample, even if the sample was statistically representative of the whole.
7. While screening for inclusion based on abstracts and titles was carried out by two people, a single author conducted full-text screening and data extraction on the 38 publications randomly sampled to create the dataset.

Future reviews featuring more databases searched, a simpler disease taxonomy, a longer time horizon, and complete coding of the entire dataset of publications eligible for inclusion, would make a useful complement to the novel dataset presented here. This novel dataset could also be used to inform the scope of such reviews.

## Conclusion

Cross-sectional prevalence studies are alive and well in sSA. A consistent annual volume of eligible studies was published in the years covered by this scoping review. Studies represented places all over the geographic area included in the review. A wide range of disease categories and individual diseases were measured by investigators. Prevalence was measured in many populations including infants, children, adults, antenatal and postnatal women, as well as specialty sub-populations. These studies took place both in basic urban and rural locales, but also in specialized places such as refugee camps and workplaces. This variation was present even within just a subset of the entire dataset.

Results from the sample show that funded studies are likely slower to complete and larger by sample size. Funding sources are the Global North, Global South, and from the investigators themselves. Use of a local university or hospital ethical review board appears to speed up a cross-sectional prevalence study as opposed to using a governmental ethical review board. While there is a large breadth across many dimensions, it seems certain countries—our sample found Ethiopia particularly—perform more of these studies than others. Certain diseases—malaria and intestinal parasitic infection—are more common than others in our sample. The studies are not fast: their duration to implement is measured in years not months.

With a strong set of benefits in prospective cross-sectional prevalence studies, and little structured funding or tools for them, it is likely more can be gained from systematic investment in this type of research. It will be years before digital health data and electronic medical records for diseases beyond PEPFAR-sponsored HIV are ubiquitous. Until that time, taking traditional, active field measurements at a point in time will be needed. It seems likely lessons can be learned about how to be faster or more efficient as well as more collaborative in these studies. The downstream effects, from understanding health to helping trials be more informative, could be large.

## Supporting information

Supplemental File 1 - Search Strategy

Supplemental File 2 - Random Sample Data

Supplemental File 3 - SCR Checklist

Supplemental File 4 - publications

## Author’s Contributions

SD: Conceptualization, Writing-Original Draft Preparation, Investigation, Methodology, Data Curation, Project Administration, Supervision, Funding Acquisition

CM: Software, Formal Analysis, Visualization, Writing-Review & Editing

PQ: Visualization

TN: Validation, Writing-Review & Editing

## Acknowledgements

This paper would not have been possible without the exceptional support of Dr. Belinda Burford.

## Ethics and Consent for Publication

This scoping review does not require ethical approval; it does not report on or involve the use of any animal or human data or tissue. Consent for publication is non-applicable.

## Data Availability

The data used in this scoping review is available at two sources. First, for the full dataset—the 363 open-access publications—, please see the Supplementary Materials for a list of citations. Second, for the coded random sample, 38 publications, see the Supplementary Materials for the detailed data.

