## Supplemental File 1 - Search Strategy for "Recent cross-sectional prevalence studies in sub-Saharan Africa for communicable, maternal, neonatal, and nutritional diseases and conditions: a scoping review"

S1 - PubMed Search Strategy

Originally returned as of 6/30/23: 868

(("Africa South of the Sahara"[tw] OR “Comoros”[tw] OR “Madagascar”[tw] OR “Mauritius”[tw] OR “Seychelles”[tw] OR "central africa"[tw] OR "southern africa"[tw] OR "western africa"[tw] OR "eastern africa"[tw] OR ("Burkina Faso"[tw] OR "Burkinabe"[tw]) OR "Cameroon*"[tw] OR ("Central African Republic"[tw] OR "Central African?"[tw]) OR ("Chad"[tw] OR “Chadian”[tw]) OR ("Republic of the Congo"[tw] OR "Congolese"[tw]) OR ("Democratic Republic of the Congo"[tw] OR “Democratic Republic of Congo”[tw] OR “DR Congo”[tw]) OR ("Equatorial Guinea*"[tw] OR "Equatoguinean?"[tw]) OR "Gabon*"[tw] OR (“Sao Tome and Principe”[tw] OR "Sao Tome*"[tw]) OR ("Burundi*"[tw] OR "Barundi"[tw]) OR ("Comoros"[tw] OR “Comorian”[tw]) OR "Djibouti*"[tw] OR "Eritrea*"[tw] OR "Ethiopia*"[tw] OR "Kenya*"[tw] OR "Madagas*"[tw] OR "Mauriti*"[tw] OR "Rwand*"[tw] OR "Seychell*"[tw] OR "Somali*"[tw] OR "South Sudan*"[tw] OR "Sudan*"[tw] OR "Tanzania*"[tw] OR "Uganda*"[tw] OR "Mauritania*"[tw] OR "Angola*" [tw] OR ("Botswana"[tw] OR "Batswana"[tw] OR "Motswana"[tw]) OR ("Eswatini"[tw] OR "Swazi*"[tw]) OR ("Lesotho"[tw] OR "Basotho"[tw] OR "Mosotho"[tw]) OR "Malawi*"[tw] OR "Mozambi*"[tw] OR "Namibia*"[tw] OR "South Africa*"[tw] OR "Zambia*"[tw] OR "Zimbabwe*"[tw] OR “Benin*"[tw] OR ("Cabo Verde*"[tw] OR "Cape Verde*"[tw]) OR (“Cote d’Ivoire”[tw] OR "Ivory Coast"[tw] OR "Ivorian*"[tw]) OR "Gambia*"[tw] OR "Ghana*"[tw] OR ("Guinea*"[tw] NOT (“New Guinea*”[tw] OR “Guinea worm”[tw])) OR ("Guinea-Bissau*"[tw] OR "Bissau-Guinean?"[tw]) OR "Liberia*"[tw] OR ("Mali"[tw] OR “Malian?”[tw]) OR ("Niger"[tw] OR “Nigerien?”[tw]) OR "Nigeria*"[tw] OR "Senegal*"[tw] OR "Sierra Leone*"[tw] OR "Togo*"[tw] OR “Tajikstan”[tw] OR “Yemen”[tw] OR “India”[tw] OR “Pakistan”[tw] OR “Afghanistan”[tw] OR “Myanmar”[tw] OR “Cambodia”[tw] OR “Timor-Leste”[tw] OR “Uzbekistan”[tw] OR “Kyrgyzstan”[tw])
AND
((“HIV Infections”[mh] OR "Sexually Transmitted Diseases"[mh] OR “Syphilis”[mh] OR “Chlamydia Infections”[mh] OR “Gonorrhea”[mh] OR “Trichomonas Vaginitis”[mh] OR “Herpes Genitalis”[mh])
OR
(“Respiratory Tract Infections”[mh] OR "Tuberculosis"[mh] OR “Latent Tuberculosis”[mh] OR “Tuberculosis, Multidrug-Resistant”[mh] OR “Extensively Drug-Resistant Tuberculosis”[mh] OR “Otitis media”[mh])
OR
(“Gastroenteritis”[mh] OR “Dysentery”[mh] OR “Typhoid Fever”[mh] OR “Paratyphoid Fever”[mh] OR “Salmonella Infections”[mh])
OR
(“Vector Borne Diseases”[mh] OR “Parasitic Diseases”[mh] OR “Waterborne Diseases”[mh] OR “Zoonoses”[mh] OR “Virus Diseases”[mh] OR “Malaria”[mh] OR “Chagas Disease”[mh] OR “Leishmaniasis, Cutaneous”[mh] OR “Leishmaniasis, Mucocutaneous”[mh] OR “Leishmaniasis, Visceral”[mh] OR “Trypanosomiasis, African”[mh] OR “Schistosomiasis”[mh] OR “Cysticercosis”[mh] OR “Echinococcosis”[mh] OR “Filariasis”[mh] OR “Onchocerciasis”[mh] OR “Trachoma”[mh] OR “Dengue”[mh] OR “Yellow Fever”[mh] OR “Rabies”[mh] OR “Ascariasis”[mh] OR “Trichuriasis”[mh] OR “Hookworm Infections”[mh] OR “Trematode Infections”[mh] OR “Leprosy”[mh] OR “Hemorrhagic Fever, Ebola”[mh] OR “Zika Virus Infection”[mh] OR “Dracunculiasis”[mh])
OR

(“Infections”[mh] OR “Meningitis”[mh] OR “Encephalitis”[mh] OR “Diphtheria”[mh] OR "Whooping Cough"[mh] OR “Tetanus”[mh] OR “Measles”[mh] OR “Varicella Zoster Virus Infection”[mh] OR “Hepatitis A”[mh] OR “Hepatitis B”[mh:noexp] OR “Hepatitis C”[mh:noexp] OR “Hepatitis E”[mh])
OR
(“Pregnancy Complications”[mh] OR “Postpartum Hemorrhage”[mh] OR “Pregnancy Complications, Infectious”[mh] OR “Hypertension, Pregnancy-Induced”[mh] OR “Dystocia”[mh] OR “Uterine Rupture”[mh] OR “Abortion, Spontaneous”[mh] OR “Pregnancy, Ectopic”[mh] OR “Maternal Death”[mh] OR “Infant, Newborn, Diseases”[mh] OR “Premature Birth”[mh] OR “Asphyxia Neonatorum”[mh] OR ("Hypoxia, Brain"[mh] AND "Infant, Newborn"[mh]) OR “Neonatal Sepsis”[mh] OR “Jaundice, Neonatal”[mh] OR “Erythroblastosis, Fetal”[mh])
OR
(“Malnutrition”[mh] OR “Vitamin A Deficiency”[mh] OR “Protein-Energy Malnutrition”[mh] OR “Iron Deficiencies”[mh] OR “Iodine/deficiency”[mh])
NOT
(“COVID-19”[mh] OR “cattle”[mh] OR “retrospective”[mh] OR “India”[ti] OR “Pakistan”[ti])))
AND
(“cross-sectional”[tw] OR “cross sectional”[tw])
AND
(“prevalence”[ti])
AND
(2019/06/01:2023/06/01 [dp])
