## Supplemental File 2 - Random Sample Data for "Recent cross-sectional prevalence studies in sub-Saharan Africa for communicable, maternal, neonatal, and nutritional diseases and conditions: a scoping review"

| Usable Count | Authors | Title of article | Text:Duration of Collection | Data Collection (Days Rounded) | Start Date of Data Collection | Data Collection Start Date to Publication Date Duration | Disease or condition | Target population | Disease or Condition Type | Target population type | Part of a randomized controlled trial (Y/N) |
|---|---|---|---|---|---|---|---|---|---|---|---|
| 1 | Yideg Yitbarek G and Ayele | Prevalence of Syphilis amor | Nov 2018 to | 90 | 11/1/2018 | 225 | syphilis | pregnant women | pregnancy | pregnant women | N |
| 2 | Tombang AN and Ambe NF | Prevalence and risk factors | April 10th to | 80 | 4/10/2018 | 497 | cryptosporidiosis | children, 0-5 | intestinal parasitic infection | children | N |
| 3 | Tilahun A and Yimer M and | Prevalence of asymptomati | Feb to March | 60 | 2/1/2019 | 445 | malaria | pregnant women | malaria | pregnant women | N |
| 4 | Sebsbie A and Minda A and | Co-existence of overweight, | May to June | 60 | 5/1/2021 | 424 | stunting | 6-59 months of age | malnutrition | children | N |
| 5 | Ngong IN and Fru-Cho J an | Prevalence, antimicrobial su | Aug to Nove | 120 | 8/1/2017 | 1,525 | urinary tract infection | pregnant women | bacterial infection | pregnant women | N |
| 6 | Carod JF and Mauny F and | Hyperendemicity of cysticer | Feb to March | 60 | 2/1/2007 | 5,356 | cysticercosis | school children | NTD | children | N |
| 7 | Ahmed A and Mulatu K and | Prevalence of malaria and a | Octo to Nov | 60 | 10/1/2019 | 507 | malaria | under 5 refugees | malaria | children | N |
| 8 | Mremi A and Rwenyagila D | Prevalence of post-partum | Oct to Dec 19 | 90 | 10/1/2019 | 856 | post-partum anemia | postnatal women | of pregnancy | postnatal women | N |
| 9 | Achaw B and Tesfa H and Z | Sero-prevalence of Toxopla | Feb to May 1 | 120 | 2/1/2018 | 518 | toxoplasma gondii | psychiatric outpatients | NTD | other | N |
| 10 | Reda G and Yemane D and | Prevalence and associated f | March of 20: | 30 | 3/1/2018 | 776 | trachoma | 1-9 yo | NTD | children | N |
| 11 | Mosha JF and Lukole E and | Risk factors for malaria infe | Sept to Oct 2 | 60 | 9/1/2014 | 2,180 | malaria | children | malaria | children | N |
| 12 | Asa BF and Shintouo CM a | Prevalence, correlates of un | April, May 19 | 60 | 4/1/2019 | 1,347 | malnutrition, intestinal parisitic in | children 1-5 | intestinal parasitic infection | children | N |
| 13 | Ahmed RH and Yussuf AA a | Anemia among pregnant w | Feb to July 20 | 180 | 2/1/2020 | 682 | anemia | pregnant women | of pregnancy | pregnant women | N |
| 14 | Okidi L and Ongeng D and | Disparity in prevalence and | Oct to Dec 19 | 90 | 10/1/2019 | 972 | malnutrition | kids 6-59 months | intestinal parasitic infection | children | N |
| 15 | Mabunda N and Augusto C | Nucleic acid testing identifie | Nov 14 to Oc | 360 | 11/1/2014 | 2,735 | HIV, HBV, HCV | adults < 65 yo | virus | adults | N |
| 16 | Ahmed ME and Abdalla SS | Prevalence of cystic echinoc | Nov 17 to Ap | 180 | 11/1/2017 | 1,053 | cystic echinococcosis | all (not clear lower bound) | NTD | adults | N |
| 17 | Budge S and Barnett M and | Risk factors and transmissio | June, 2019 | 30 | 6/1/2019 | 342 | malnutrition, intestinal parisitic in | infants 10-18 months | intestinal parasitic infection | infants | N |
| 18 | Mehari S and Zerfu B and D | Prevalence and risk factors | Feb to May 2 | 120 | 2/1/2019 | 952 | malaria & human brucellosis | adult, 2 yo+ | malaria | adults | N |
| 19 | Nwaneli EI and Eguonu I ar | Malaria prevalence and its s | June 16 to Ja | 240 | 6/1/2016 | 1,461 | malaria | children, 6 mo -17 yo | malaria | children | N |
| 20 | Kefale A and Daka K and Al | Prevalence of HIV seroposit | Feb 18 to Jur | 130 | 2/18/2021 | 725 | HIV | family members of HIV+ people | virus | special | N |
| 21 | Ejerssa AW and Gadisa DA | Prevalence of bacterial urop | Nov 17 to Jar | 90 | 11/1/2017 | 1,375 | uropathogens | pregnant women | of pregnancy | pregnant women | N |
| 22 | Amare A and Eshetie S and | High prevalence of fecal car | Feb to June 2 | 150 | 2/1/2021 | 409 | ESBL-EC, ESBL-KP | food handlers at uni | bacterial infection | special | N |
| 23 | Cote CM and Goel V and M | Malaria prevalence and lon | Jan 8 to Mar | 90 | 1/8/2020 | 544 | malaria | children 2-10 yo | malaria | children | N |
| 24 | Yaya I and Boyer V and Ehl | Heterogeneity in the Preval | Aug 17 to De | 510 | 8/1/2017 | 1,298 | HPV | men who have sex with men | virus | other | N |
| 25 | Nwankwo HC and Habtu M | Prevalence and factors asso | Aug-Sep 20 | 60 | 8/1/2020 | 856 | preterm birth | postnatal women, 18-49 yo | of pregnancy | postnatal women | N |
| 26 | Ndizeye Z and Vanden Bro | Prevalence and genotype-sp | Mar-May16 | 90 | 3/1/2016 | 1,211 | HPV | women attending one of 2 clini | virus | women | N |
| 27 | Abbas AA and Ali HAA and | Prevalence and risk factors | May to July 1 | 90 | 5/1/2019 | 519 | Hantavirus | 20-80 yo adults | NTD | adults | N |
| 28 | Macicame I and Bhatt N an | HIV prevalence and risk beh | Nov 13 to No | 390 | 11/1/2013 | 2,146 | HIV | enrollees to RCT vax prep sessi | virus | special | Y |
| 29 | Ali AH and Reda DY and Or | Prevalence and antimicrobi | May to Oct 2 | 180 | 5/1/2020 | 635 | urinary tract infection | pregnant women | pregnancy | pregnant women | N |
| 30 | Tsegaye B and Yoseph A ar | Prevalence and factors asso | Jan 1 to 20 1 | 30 | 1/1/2018 | 751 | intestinal parasitic infection | children, 6-59 mos | intestinal parasitic infection | children | N |
| 31 | Kharsany ABM and McKinn | Population prevalence of se | June 14 to Ju | 360 | 6/1/2014 | 2,284 | sexually transmitted infections | adults, 15-49 yo | sexually transmitted infections | adults | N |
| 32 | Chea N and Tegene Y and A | Prevalence of undernutritio | April 30 to M | 30 | 4/1/2019 | 1,500 | malnutrition | pregnant women | malnutrition | pregnant women | N |
| 33 | Sisay A and Lemma B | Assessment on the prevaler | 16-May | 30 | 5/1/2016 | 1,170 | intestinal parasitic infection | school children aged 7-25 (whic | intestinal parasitic infection | children | N |
| 34 | Geda N and Beyene T and | Prevalence of Cryptococcal | Sep 1 to Oct | 60 | 9/1/2017 | 825 | cryptococcis | HIV patients over 18 | fungal infection | adults | N |
| 35 | Oyegue-Liabagui SL and Nd | Molecular prevalence of int | Nov 16 to Au | 300 | 11/1/2016 | 1,291 | intestinal parasitic infection | children 2-180 months (0-14 yo | intestinal parasitic infection | children | N |
| 36 | Shiferaw MB and Sinishaw | Prevalence of active tuberc | Oct 1 18 to Ja | 120 | 10/1/2018 | 984 | tuberculosis | healthcare workers | bacterial infection | adults | N |
| 37 | Kayondo SP and Byamugisl | Prevalence of hepatitis B vir | Dec 18 - Feb | 90 | 12/1/2018 | 521 | HBV | pregnant women | virus | pregnant women | N |
| 38 | Hailegebriel T and Nibret E | Prevalence, intensity and as | Feb to Apr 2 | 90 | 2/1/2021 | 268 | schistosoma mansoni | schoolchildren | intestinal parasitic infection | children | N |

| Usable Count | Country | Site(s) | Prevalence Data Collection Method | Prevalence Data Collection Method Type | Date Study Article Received by Journal | Date Study Article Received by Journal | Funder (Y/N) | Funder | Funder Type | Funding Source Geography | Ethics Reviewer(s) | Ethics IRB Type | Ethics Review Public vs Non-public | Sample Size | Count of non-preganancy studies for utility of rows | Data Collection Start Date to Article Receipt Date Duration |
|---|---|---|---|---|---|---|---|---|---|---|---|---|---|---|---|---|
| 1 | Ethiopia | 2 health centers, Muja He | Institution-Hospital | Institution | 3/11/2019 | 6/14/2019 | N | none | None | Unfunded | Sede Muja woreda admin | public | public | 217 | | 130 |
| 2 | Cameroon | Limbe Regional Hosp. Ped | Institution-Hospital | Institution | 2/13/2019 | 8/20/2019 | N | none | None | Unfunded | Univ of Buea IRB | local uni | non-public | 112 | 1 | 309 |
| 3 | Ethiopia | 2 sites: Jawi Hospital & Ja | Institution-Hospital | Institution | 10/17/2019 | 4/21/2020 | Y | Bahir Dar Univ (p | University | Global South | Bahir Dar Univ Eth Rev Bd | local uni | non-public | 331 | | 258 |
| 4 | Ethiopia | 29 health centers across A | Institution-Hospital | Institution | 1/18/2022 | 6/29/2022 | N | none | None | Unfunded | Addis Ababa Health Bure | public | public | 422 | 2 | 262 |
| 5 | Cameroon | 6 hospitals in Bueau | Institution-Hospital | Institution | 1/20/2021 | 10/4/2021 | N | none | None | Unfunded | Univ of Buea IRB | local uni | non-public | 284 | | 1,268 |
| 6 | Madagascar | 7 cities;2-3 schools in eac | Institution-School | Institution | 8/28/2020 | 10/1/2021 | Y | Institut Pasteur d | Private | Global North | National Ethical Committe | public | public | 1,751 | 3 | 4,957 |
| 7 | Ethiopia | Sherkole Refugee Camp | Community-Public | Community | 8/17/2020 | 2/19/2021 | N | none | None | Unfunded | Bahir Dar Univ Eth Rev Bd | local uni | non-public | 356 | 4 | 321 |
| 8 | Tanzania | 3 urban health centers | Institution-Hospital | Institution | 4/5/2021 | 2/3/2022 | N | none | None | Unfunded | Muhimbili Univ Res Comr | local uni | non-public | 424 | 5 | 552 |
| 9 | Ethiopia | Univ of Gondar Hospital | Institution-Hospital | Institution | 9/29/2018 | 7/4/2019 | Y | Univ of Gondar (p | University | Global South | Univ of Gondar Ethical Re | local uni | non-public | 304 | 6 | 240 |
| 10 | Ethiopia | 7 kebeles out of the 24 in | Community-Households | Community | 1/6/2020 | 4/15/2020 | Y | Mekelle Univ (full | University | Global South | Mekelle Univ Health Eth R | local uni | non-public | 502 | 7 | 676 |
| 11 | Tanzania | a district, cluster | Community-Households | Community | 4/2/2020 | 8/20/2020 | Y | DFID,MRC,Wellco | Private | Global North | Kilimanjaro Medical Colle | public | public | 3,456 | 8 | 2,040 |
| 12 | Cameroon | 14 villages in thick forest | Community-Households | Community | 2/4/2022 | 12/8/2022 | Y | Center for Intl Fo | Public | Global North | Cameroon Ethics Commit | public | public | 442 | 9 | 1,040 |
| 13 | Somalia | 3 districts of refugees, Eve | Community-Households | Community | 3/2/2021 | 12/14/2021 | N | none | None | Unfunded | Jazeera Univ IRB | local uni | non-public | 383 | | 395 |
| 14 | Uganda | 3 districts, cluster | Community-Households | Community | 8/21/2021 | 5/30/2022 | Y | COE in Sustainabl | University | Global South | Gulu University Res Ethics | local uni | non-public | 183 | 10 | 690 |
| 15 | Mozambique | 2 hosp., blood banks in th | Institution-Hospital | Institution | 8/13/2021 | 4/28/2022 | Y | Flemish Gov't (in | Public | Global North | Mozambique National He | public | public | 2,783 | 11 | 2,477 |
| 16 | Sudan | 3 major cities within Khar | Community-Households | Community | 4/15/2020 | 9/19/2020 | N | none | None | Unfunded | Al-Neelain Univ, Khartoun | local uni | non-public | 305 | 12 | 896 |
| 17 | Ethiopia | 2 neighborhoods | Community-Households | Community | 12/9/2019 | 5/8/2020 | Y | Cranfield Univ UK | Private | Global North | Hawassa Univ IRB, Cranfi | other | non-public | 35 | 13 | 191 |
| 18 | Ethiopia | 1 hosp, 1 health ctr | Institution-Hospital | Institution | 10/16/2020 | 9/10/2021 | N | none | None | Unfunded | Addis Ababa Univ Hlth Re | local uni | non-public | 444 | 14 | 623 |
| 19 | Nigeria | 1 hosp (NAUTH) | Institution-Hospital | Institution | 7/24/2019 | 6/1/2020 | N | none | None | Unfunded | NAUTH Hosp Res & Ethics | hospital IRB | non-public | 382 | 15 | 1,148 |
| 20 | Ethiopia | 3 ART clinics all in hospita | Institution-Hospital | Institution | 3/2/2022 | 2/13/2023 | N | none | None | Unfunded | Wolaita Soto Univ | local uni | non-public | 623 | 16 | 377 |
| 21 | Ethiopia | antenatal care clinic locat | Institution-Hospital | Institution | 3/4/2021 | 8/7/2021 | Y | Herari Region He | Public | Global South | Addis Ababa Univ Hlth Re | local uni | non-public | 200 | | 1,219 |
| 22 | Ethiopia | 6 cafeterias at Univ of Go | Community-Public | Community | 12/5/2021 | 3/17/2022 | N | none | None | Unfunded | Univ of Gondar Ethical Re | local uni | non-public | 290 | 17 | 307 |
| 23 | Uganda | 35 villages, 60 HH per | Community-Households | Community | 4/9/2021 | 7/5/2021 | Y | US NIH NIAID | Public | Global North | UNC Chapel Hill IRB, Ugan | public | public | 2,170 | 18 | 457 |
| 24 | 4 countries i | ART-type clinics | Institution-Hospital | Institution | 11/26/2020 | 2/19/2021 | Y | Sidaction (a chari | Private | Global North | national ethics committee | public | public | 667 | 19 | 1,213 |
| 25 | Rwanda | Kabutare Hosp | Institution-Hospital | Institution | 3/4/2022 | 12/5/2022 | N | none | None | Unfunded | Mt Kenya Univ | local uni | non-public | 240 | 20 | 580 |
| 26 | Burundi | one HIV clinic, one fam pl | Institution-Hospital | Institution | 11/22/2018 | 6/25/2019 | Y | IUC-VLIRUOS/ Un | Private | Global North | Burundian Natl Ethics Cor | public | public | 300 | 21 | 996 |
| 27 | Sudan | DR Salma Dialysis Ctr | Institution-Hospital | Institution | 5/18/2020 | 10/1/2020 | N | none | None | Unfunded | Ministry of Higher Ed & S | public | public | 91 | 22 | 383 |
| 28 | Mozambique | commercial research ctr c | Community-Public | Community | 6/15/2018 | 9/17/2019 | Y | US DOD & Jackso | Public | Global North | Natl Health Bioethics Moz | public | public | 1,125 | 23 | 1,687 |
| 29 | Somalia | Hargeisa Grp Hosp | Institution-Hospital | Institution | 10/6/2021 | 1/26/2022 | N | none | None | Unfunded | Hawassa Univ IRB | local uni | non-public | 422 | | 523 |
| 30 | Ethiopia | Household 1st, then healt | Community-Households | Community | 9/19/2019 | 1/22/2020 | N | none | None | Unfunded | Hawassa Univ IRB | local uni | non-public | 622 | 24 | 626 |
| 31 | South Africa | households | Community-Households | Community | 5/3/2020 | 9/1/2020 | Y | US CDC Pepfar | Public | Global North | Uni KwaZulu-Natal & US C | public | public | 9,812 | 25 | 2,163 |
| 32 | Ethiopia | households in randomly s | Community-Households | Community | 9/8/2022 | 5/10/2023 | Y | Hawassa Univ | University | Global South | Hawassa Univ IRB | local uni | non-public | 550 | | 1,256 |
| 33 | Ethiopia | Bochesa Elementary Scho | Institution-School | Institution | 4/3/2019 | 7/15/2019 | N | none | None | Unfunded | Woreda Health Office | public | public | 384 | 26 | 1,067 |
| 34 | Ethiopia | 2 hospitals | Institution-Hospital | Institution | 7/27/2019 | 12/5/2019 | N | none | None | Unfunded | Wollega Univ | local uni | non-public | 186 | 27 | 694 |
| 35 | Gabon | 2 hospitals | Institution-Hospital | Institution | 12/27/2019 | 5/15/2020 | Y | Total Gabon | Private | Global South | Gabonese Natl Ethics Con | public | public | 100 | 28 | 1,151 |
| 36 | Ethiopia | 2 workplaces (at hospitals | Community-Public | Community | 2/3/2021 | 6/11/2021 | Y | USAID | Public | Global North | Amhara Public Health Ins | public | public | 580 | 29 | 856 |
| 37 | Uganda | 1 antenatal clinic at 1 hos | Institution-Hospital | Institution | 1/27/2020 | 5/5/2020 | Y | African Academy | Public | Global South | Makerere Univ & Uganda | public | public | 340 | | 422 |
| 38 | Ethiopia | 8 schools | Institution-School | Institution | 8/26/2021 | 10/27/2021 | Y | Bahir Dar Univ | University | Global South | Bahir Dar Univ Eth Rev Bd | local uni | non-public | 681 | 30 | 206 |

S2 - Fully coded random sample of dataset

### Recent cross-sectional prevalence studies in sub-Saharan Africa for communicable, maternal, neonatal, and nutritional diseases and conditions: a scoping review
