## Supplemental File 3 - SCR Checklist for "Recent cross-sectional prevalence studies in sub-Saharan Africa for communicable, maternal, neonatal, and nutritional diseases and conditions: a scoping review"

S3 – PRISMA-ScR Checklist

| Section | Subsection | Item | Item Description | Where it appears (page) |
|---|---|---|---|---|
| Title | [this cell deliberately left blank in compliance with PRISMA-ScR Checklist, Expanded version] | 1 | Identify the report as a scoping review. | 1 |
| Abstract | Structured Summary | 2 | Provide a structured summary that includes (as applicable) background, objectives, eligibility criteria, sources of evidence, charting/analysis methods, results, and conclusions that relate to the review questions and objectives. | 1 |
| Introduction | Rationale | 3 | Describe the rationale for the review in the context of what is already known. Explain why the review questions/objectives lend themselves to a scoping review approach. | 3 |
| | Objectives | 4 | Provide an explicit statement of the questions and objectives being addressed with reference to their key elements (e.g., population or participants, concepts, and context) or other relevant key elements used to conceptualize the review questions and/or objectives. | 5 |
| Methods | Protocol and registration | 5 | Indicate whether a review protocol exists; state if and where it can be accessed (e.g., a Web address); and if available, provide registration information, including the registration number. | 6 |
| | Eligibility criteria | 6 | Specify characteristics of the sources of evidence used as eligibility criteria (e.g., years considered, language, and publication status), and provide a rationale. | 6 |
| | Information sources | 7 | Describe all information sources in the search (e.g., databases with dates of coverage and contact with authors to identify additional sources), as well as the date the most recent search was executed. | 6 |
| | Search | 8 | Present the full electronic search strategy for at least 1 database, including any limits used, such that it could be repeated. | 28 |
| | Selection of sources of evidence | 9 | State the process for selecting sources of evidence (i.e., screening and eligibility) included in the scoping review. | 8 |
| | Data charting/analysis process | 10 | Describe the methods of charting/analyzing data from the included sources of evidence (e.g., calibrated forms or forms that have been tested by the team before their use, and whether data charting/analysis was done independently or in duplicate) and any processes for obtaining and confirming data from investigators. | 11 |
| | Data items | 11 | List and define all variables for which data were sought and any assumptions and simplifications made. | 10 |
| | Critical appraisal of individual sources of evidence | 12 | If done, provide a rationale for conducting a critical appraisal of included sources of evidence; describe the methods used and how this information was used in any data synthesis (if appropriate). | 12 |
| | Summary measures | 13 | Not applicable for scoping reviews | n/a |
| | Synthesis of results | 14 | Describe the methods of handling and summarizing the data that were charted/analyzed. | 15 |
| | Risk of bias across studies | 15 | Not applicable for scoping reviews | n/a |
| | Additional analyses | 16 | Not applicable for scoping reviews | n/a |
| Results | Selection of sources of evidence | 17 | Give numbers of sources of evidence screened, assessed for eligibility, and included in the review, with reasons for exclusions at each stage, ideally using a flow diagram. | 13 |
| | Characteristics of sources of evidence | 18 | For each source of evidence, present characteristics for which data were charted/analyzed and provide the citations. | 14 |
| | Critical appraisal within sources of evidence | 19 | If done, present data on critical appraisal of included sources of evidence (see item 12). | 12 |
| | Results of individual sources of evidence | 20 | For each included source of evidence, present the relevant data that were charted/analyzed that relate to the review questions and objectives. | 30 |
| | Synthesis of results | 21 | Summarize and/or present the charting/analysis results as they relate to the review questions and objectives. | 15 |
| | Risk of bias across studies | 22 | Not applicable for scoping reviews | n/a |
| | Additional analyses | 23 | Not applicable for scoping reviews | n/a |
| Discussion | Summary of evidence | 24 | Summarize the main results (including an overview of concepts, themes, and types of evidence available), link to the review questions and objectives, and consider the relevance to key groups. | 22 |
| | Limitations | 25 | Discuss the limitations of the scoping review process | 22 |
| | Conclusion | 26 | Provide a general interpretation of the results with respect to the review questions and objectives, as well as potential implications and/or next steps. | 23 |
| Funding | [this cell deliberately left blank in compliance with PRISMA-ScR Checklist, Expanded version] | 27 | Describe sources of funding for the included sources of evidence, as well as sources of funding for the scoping review. Describe the role of the funders of the scoping review. | 25 |
