## Supplemental File 4 - publications for "Recent cross-sectional prevalence studies in sub-Saharan Africa for communicable, maternal, neonatal, and nutritional diseases and conditions: a scoping review"

S4 - Dataset citations


1. Yideg Yitbarek G, Ayele BA. Prevalence of syphilis among pregnant women attending antenatal care clinic, Sede Muja District, South Gondar, Northwest Ethiopia. Journal of pregnancy. 2019 Jul 14;2019.
2. Tombang AN, Ambe NF, Bobga TP, Nkfusai CN, Collins NM, Ngwa SB, Diengou NH, Cumber SN. Prevalence and risk factors associated with cryptosporidiosis among children within the ages 0–5 years attending the Limbe regional hospital, southwest region, Cameroon. BMC Public Health. 2019 Dec;19(1):1-0.
3. Tilahun A, Yimer M, Gelaye W, Tegegne B. Prevalence of asymptomatic Plasmodium species infection and associated factors among pregnant women attending antenatal care at Fendeka town health facilities, Jawi District, North west Ethiopia: a cross-sectional study. PLoS One. 2020 Apr 21;15(4):e0231477.
4. Sebsbie A, Minda A, Ahmed S. Co-existence of overweight/obesity and stunting: it's prevalence and associated factors among under-five children in Addis Ababa, Ethiopia. BMC pediatrics. 2022 Jun 29;22(1):377.
5. Ngong IN, Fru-Cho J, Yung MA, Akoachere JF. Prevalence, antimicrobial susceptibility pattern and associated risk factors for urinary tract infections in pregnant women attending ANC in some integrated health centers in the Buea Health District. BMC Pregnancy and Childbirth. 2021 Dec;21:1-0.
6. Carod JF, Mauny F, Parmentier AL, Desmarets M, Rakotondrazaka M, Brembilla A, Dermauw V, Razafimahefa J, Ramahefarisoa RM, Andriantseheno M, Bailly S. Hyperendemicity of cysticercosis in Madagascar: Novel insights from school children population-based antigen prevalence study. Plos one. 2021 Oct 1;16(10):e0258035.
7. Ahmed A, Mulatu K, Elfu B. Prevalence of malaria and associated factors among under-five children in Sherkole refugee camp, Benishangul-Gumuz region, Ethiopia. A cross-sectional study. PloS one. 2021 Feb 19;16(2):e0246895.
8. Mremi A, Rwenyagila D, Mlay J. Prevalence of post-partum anemia and associated factors among women attending public primary health care facilities: An institutional based cross-sectional study. Plos one. 2022 Feb 3;17(2):e0263501.
9. Achaw B, Tesfa H, Zeleke AJ, Worku L, Addisu A, Yigzaw N, Tegegne Y. Sero-prevalence of Toxoplasma gondii and associated risk factors among psychiatric outpatients attending University of Gondar Hospital, Northwest Ethiopia. BMC Infectious Diseases. 2019 Dec;19(1):1-8.
10. Reda G, Yemane D, Gebreyesus A. Prevalence and associated factors of active trachoma among 1–9 years old children in Deguatemben, Tigray, Ethiopia, 2018: community cross-sectional study. BMC ophthalmology. 2020 Dec;20(1):1-9.
11. Mosha JF, Lukole E, Charlwood JD, Wright A, Rowland M, Bullock O, Manjurano A, Kisinza W, Mosha FW, Kleinschmidt I, Protopopoff N. Risk factors for malaria infection prevalence and household vector density between mass distribution campaigns of long-lasting insecticidal nets in North-western Tanzania. Malaria journal. 2020 Dec;19:1-1.
12. Asa BF, Shintouo CM, Shey RA, Afoumbom MT, Siekeh N, Yoah A, Kah E, Ickowitz A, Tata CY, Asongalem E, Ghogomu SM. Prevalence, correlates of undernutrition and intestinal parasitic infection among children below 5 years living in the forest community of Ndelele, East Region of Cameroon: A cross-sectional assessment. Plos one. 2022 Dec 8;17(12):e0278333.
13. Ahmed RH, Yussuf AA, Ali AA, Iyow SN, Abdulahi M, Mohamed LM, Mohamud MH. Anemia among pregnant women in internally displaced camps in Mogadishu, Somalia: a cross-sectional study on prevalence, severity and associated risk factors. BMC Pregnancy and Childbirth. 2021 Dec;21(1):1-9.

14. Okidi L, Ongeng D, Muliro PS, Matofari JW. Disparity in prevalence and predictors of undernutrition in children under five among agricultural, pastoral, and agro-pastoral ecological zones of Karamoja sub-region, Uganda: a cross sectional study. BMC pediatrics. 2022 May 30;22(1):316.
15. Mabunda N, Augusto O, Zicai AF, Duajá A, Oficiano S, Ismael N, Vubil A, Mussá T, Moraes M, Jani I. Nucleic acid testing identifies high prevalence of blood borne viruses among approved blood donors in Mozambique. PLoS One. 2022 Apr 28;17(4):e0267472.
16. Ahmed ME, Abdalla SS, Adam IA, Grobusch MP, Aradaib IE. Prevalence of cystic echinococcosis and associated risk factors among humans in Khartoum State, Central Sudan. International Health. 2021 Jul;13(4):327-33.
17. Budge S, Barnett M, Hutchings P, Parker A, Tyrrel S, Hassard F, Garbutt C, Moges M, Woldemedhin F, Jemal M. Risk factors and transmission pathways associated with infant Campylobacter spp. prevalence and malnutrition: a formative study in rural Ethiopia. PLoS One. 2020 May 8;15(5):e0232541.
18. Mehari S, Zerfu B, Desta K. Prevalence and risk factors of human brucellosis and malaria among patients with fever in malaria-endemic areas, attending health institutes in Awra and Gulina district, Afar Region, Ethiopia. BMC Infectious Diseases. 2021 Dec;21:1-8.
19. Nwaneli EI, Eguonu I, Ebenebe JC, Osuorah CD, Ofiaeli OC, Nri-Ezedi CA. Malaria prevalence and its sociodemographic determinants in febrile children-a hospital-based study in a developing community in South-East Nigeria. Journal of preventive medicine and hygiene. 2020 Jun;61(2):E173.
20. Kefale A, Daka K, Abebe A, Haile D, Paulos K, Sherfa A, Addis A, Gunta M, Ayza A, Wolde J. Prevalence of HIV seropositive status and associated factors among family members of index cases of antiretroviral clinical attendants in Sodo Town, Southern Ethiopia. Plos one. 2023 Feb 13;18(2):e0280571.
21. Ejerssa AW, Gadisa DA, Orjino TA. Prevalence of bacterial uropathogens and their antimicrobial susceptibility patterns among pregnant women in Eastern Ethiopia: hospital-based cross-sectional study. BMC Women's Health. 2021 Aug 7;21(1):291.
22. Amare A, Eshetie S, Kasew D, Moges F. High prevalence of fecal carriage of Extended-spectrum beta-lactamase and carbapenemase-producing Enterobacteriaceae among food handlers at the University of Gondar, Northwest Ethiopia. Plos one. 2022 Mar 17;17(3):e0264818.
23. Cote CM, Goel V, Muhindo R, Baguma E, Ntaro M, Shook-Sa BE, Reyes R, Staedke SG, Mulogo EM, Boyce RM. Malaria prevalence and long-lasting insecticidal net use in rural western Uganda: results of a cross-sectional survey conducted in an area of highly variable malaria transmission intensity. Malaria journal. 2021 Dec;20(1):1-2.
24. Yaya I, Boyer V, Ehlan PA, Coulibaly A, Agboyibor MK, Traoré I, Kouamé MJ, Maiga AK, Kotchi OR, Nyasenu YT, Maradan G. Heterogeneity in the prevalence of high-risk human papillomavirus infection in human immunodeficiency virus-negative and human immunodeficiency virus-positive men who have sex with men in West Africa. Clinical Infectious Diseases. 2021 Dec 15;73(12):2184-92.
25. Nwankwo HC, Habtu M, Rutayisire E, Kalisa R. Prevalence and factors associated with preterm birth in a rural district hospital, Rwanda. The Pan African Medical Journal. 2022;43.
26. Ndizeye Z, Vanden Broeck D, Lebelo RL, Bogers J, Benoy I, Van Geertruyden JP. Prevalence and genotype-specific distribution of human papillomavirus in Burundi according to HIV status and urban or rural residence and its implications for control. PLoS One. 2019 Jun 25;14(6):e0209303.
27. Abbas AA, Ali HA, Alagib MA, Salih HF, Elkhidir IM, El Hussein AR, Enan KA. Prevalence and risk factors of Hantavirus infection in patients undergoing hemodialysis in Khartoum, Sudan, in 2019: a cross-sectional study. Transactions of the Royal Society of Tropical Medicine and Hygiene. 2021 Jun;115(6):664-8.
28. Macicame I, Bhatt N, Matavele Chissumba R, Eller LA, Viegas E, Araújo K, Nwoga C, Li Q, Milazzo M, Hills NK, Lindan C. HIV prevalence and risk behavior among male and female adults screened for enrolment into a vaccine preparedness study in Maputo, Mozambique. PLoS One. 2019 Sep 17;14(9):e0221682.
29. Ali AH, Reda DY, Ormago MD. Prevalence and antimicrobial susceptibility pattern of urinary tract infection among pregnant women attending Hargeisa Group Hospital, Hargeisa, Somaliland. Scientific Reports. 2022 Jan 26;12(1):1419.
30. Tsegaye B, Yoseph A, Beyene H. Prevalence and factors associated with intestinal parasites among children of age 6 to 59 months in, Boricha district, South Ethiopia, in 2018. BMC pediatrics. 2020 Dec;20:1-7.
31. Kharsany AB, McKinnon LR, Lewis L, Cawood C, Khanyile D, Maseko DV, Goodman TC, Beckett S, Govender K, George G, Ayalew KA. Population prevalence of sexually transmitted infections in a high HIV

burden district in KwaZulu-Natal, South Africa: Implications for HIV epidemic control. International Journal of Infectious Diseases. 2020 Sep 1;98:130-7.
32. Chea N, Tegene Y, Astatkie A, Spigt M. Prevalence of undernutrition among pregnant women and its differences across relevant subgroups in rural Ethiopia: a community-based cross-sectional study. Journal of Health, Population and Nutrition. 2023 Dec;42(1):1-0.
33. Sisay A, Lemma B. Assessment on the prevalence and risk factors of gastrointestinal parasites on schoolchildren at Bochesa Elementary School, around Lake Zwai, Ethiopia. BMC research notes. 2019 Dec;12:1-6.
34. Geda N, Beyene T, Dabsu R, Mengist HM. Prevalence of Cryptococcal Antigenemia and associated factors among HIV/AIDS patients on second-line antiretroviral therapy at two hospitals in Western Oromia, Ethiopia. PloS one. 2019 Dec 5;14(12):e0225691.
35. Oyegue-Liabagui SL, Ndjangangoye NK, Kouna LC, Lekolo GM, Mounioko F, Kwedi Nolna S, Lekana-Douki JB. Molecular prevalence of intestinal parasites infections in children with diarrhea in Franceville, Southeast of Gabon. BMC Infectious Diseases. 2020 Dec;20:1-1.
36. Shiferaw MB, Sinishaw MA, Amare D, Alem G, Asefa D, Klinkenberg E. Prevalence of active tuberculosis disease among healthcare workers and support staff in healthcare settings of the Amhara region, Ethiopia. Plos one. 2021 Jun 11;16(6):e0253177.
37. Kayondo SP, Byamugisha JK, Ntuyo P. Prevalence of hepatitis B virus infection and associated risk factors among pregnant women attending antenatal clinic in Mulago Hospital, Uganda: a cross-sectional study. BMJ open. 2020 Jun 1;10(6):e033043.
38. Hailegebriel T, Nibret E, Munshea A, Ameha Z. Prevalence, intensity and associated risk factors of Schistosoma mansoni infections among schoolchildren around Lake Tana, northwestern Ethiopia. PLoS Neglected Tropical Diseases. 2021 Oct 27;15(10):e0009861.
39. Tarekegn M, Tekie H, Dugassa S, Wolde-Hawariat Y. Malaria prevalence and associated risk factors in Dembiya district, North-western Ethiopia. Malaria Journal. 2021 Dec;20:1-1.
40. Mulindwa J, Namulondo J, Kitibwa A, Nassuuna J, Nyangiri OA, Kimuda MP, Boobo A, Nerima B, Busingye F, Candia R, Namukuta A. High prevalence of Schistosoma mansoni infection and stunting among school age children in communities along the Albert-Nile, Northern Uganda: A cross sectional study. PLoS neglected tropical diseases. 2022 Jul 27;16(7):e0010570.
41. Zenebe MH, Mekonnen Z, Loha E, Padalko E. Prevalence, risk factors and association with delivery outcome of curable sexually transmitted infections among pregnant women in Southern Ethiopia. PloS one. 2021 Mar 24;16(3):e0248958.
42. Adeyemi OA, Itanyi IU, Ozigbu CE, Stadnick N, Tsuyuki K, Olayiwola O, Ogidi AG, Eze C, Aarons GA, Onoka CA, Ezeanolue EE. Sero-prevalence and determinants of Hepatitis B among a cohort of HIV-infected women of reproductive age in Nigeria. PloS one. 2020 Sep 17;15(9):e0236456.
43. Ayelgn K, Guadu T, Getachew A. Low prevalence of active trachoma and associated factors among children aged 1–9 years in rural communities of Metema District, Northwest Ethiopia: a community based cross-sectional study. Italian Journal of Pediatrics. 2021 Dec;47(1):1-8.
44. Tuta KE, Okesola AO, Umeokonkwo CD. The prevalence and risk factors associated with nasal methicillin-resistant Staphylococcus aureus colonization among children in a tertiary hospital in Nigeria. Ethiopian Journal of Health Sciences. 2019;29(4).
45. Tanga AT, Teshome MA, Hiko D, Fikru C, Jilo GK. Sero-prevalence of hepatitis B virus and associated factors among pregnant women in Gambella hospital, South Western Ethiopia: facility based cross-sectional study. BMC infectious diseases. 2019 Dec;19(1):1-7.
46. Mutabazi SA, Jumanne S, Mpondo BC, Mnzava DP. Prevalence of culture positive Tuberculosis and utility of a clinical diagnostic tool for the diagnosis of Tuberculosis among HIV Infected Children attending HIV/AIDS Care and Treatment in Dodoma Municipality, Central Tanzania. International Journal of Infectious Diseases. 2020 Jul 1;96:593-9.
47. Ilboudo B, Traoré I, Méda CZ, Hien A, Kinda M, Dramaix-Wilmet M, Savadogo LG, Donnen P. Prevalence and factors associated with anaemia in pregnant women in Cascades Region of Burkina Faso in 2012. Pan African Medical Journal. 2021 Apr 14;38(1).

48. Sivahikyako SA, Owaraganise A, Tibaijuka L, Agaba DC, Kayondo M, Ngonzi J, Mugisha J, Kanyesigye H. Prevalence and factors associated with severe anaemia post-caesarean section at a tertiary Hospital in Southwestern Uganda. BMC Pregnancy and Childbirth. 2021 Dec;21(1):1-8.
49. Fentahun A, Hailu T, Alemu G. Prevalence of intestinal parasites and Schistosoma mansoni and associated factors among fishermen at Lake Tana, Northwest Ethiopia. BioMed Research International. 2021 Nov 30;2021.
50. Hajare ST, Chekol Y, Chauhan NM. Assessment of prevalence of Giardia lamblia infection and its associated factors among government elementary school children from Sidama zone, SNNPR, Ethiopia. Plos one. 2022 Mar 15;17(3):e0264812.
51. Belay AS, Yehualashet SS, Abateneh DD, Kebede KM. Sero-prevalence of Hepatitis B virus surface antigen and associated factors among women of reproductive age in Bench Maji Zone, Southwest Ethiopia: Community based cross-sectional study. African health sciences. 2022 Jul 29;22(2):107-15.
52. Hassan K, Kyriakakis C, Doubell A, Van Zyl G, Claassen M, Zaharie D, Herbst P. Prevalence of cardiotropic viruses in adults with clinically suspected myocarditis in South Africa. Open Heart. 2022 Jan 1;9(1):e001942.
53. Gebrehiwet MG, Medhaniye AA, Alema HB. Prevalence and associated factors of soil transmitted helminthes among pregnant women attending antenatal care in Maytsebri primary hospital, North Ethiopia. BMC Research Notes. 2019 Dec;12(1):1-6.
54. Sadia-Kacou CA, Adja MA, Assi SB, Poinsignon A, Coulibaly JT, Ouattara AF, Remoué F, Koudou BG, Tano Y. Seasonal prevalence of Plasmodium falciparum infection and use of insecticide-treated nets among children in three agroecosystems in Aboisso, Côte d'Ivoire. Parasitology Research. 2021 Nov;120:3663-71.
55. Fondjo LA, Addai-Mensah O, Annani-Akollor ME, Quarshie JT, Boateng AA, Assafuah SE, Owiredu EW. A multicenter study of the prevalence and risk factors of malaria and anemia among pregnant women at first antenatal care visit in Ghana. PloS one. 2020 Aug 21;15(8):e0238077.
56. Shama AT, Wakuma O, Debelo S, Terefa DR, Cheme MC, Lema M, Biru B, Geta ET. Prevalence and associated factors of stunting and thinness among primary school-aged children in Gudeya Bila district, West Ethiopia: a cross-sectional study. BMJ open. 2023 May 1;13(5):e072313.
57. Jisuvei SC, Osoti A, Njeri MA. Prevalence, antimicrobial susceptibility patterns, serotypes and risk factors for group B streptococcus rectovaginal isolates among pregnant women at Kenyatta National Hospital, Kenya; a cross-sectional study. BMC infectious diseases. 2020 Dec;20(1):1-9.
58. Eyayu T, Yimer G, Workineh L, Tiruneh T, Sema M, Legese B, Almaw A, Solomon Y, Malkamu B, Chanie ES, Feleke DG. Prevalence, intensity of infection and associated risk factors of soil-transmitted helminth infections among school children at Tachgayint woreda, Northcentral Ethiopia. Plos one. 2022 Apr 8;17(4):e0266333.
59. Hailu T, Mulu W, Abera B. Prevalence and determinant factors of hookworm infection among school age children in Jawe district, NorthWest Ethiopia. African health sciences. 2019 Nov 5;19(3):2439-45.
60. Temesgen E, Belete Y, Haile K, Ali S. Prevalence of active tuberculosis and associated factors among people with chronic psychotic disorders at St. Amanuel Mental Specialized Hospital and Gergesenon Mental Rehabilitation center, Addis Ababa, Ethiopia. BMC Infectious Diseases. 2021 Dec;21(1):1-9.
61. Rambiki E, Dimba A, Ng'ambi W, Banda K, Girma B, Shight B, Lwanda L, Dambe I, Tripathy JP, Chola M, Chanda-Kapata P. The prevalence of pulmonary tuberculosis among miners from the Karonga, Rumphi, Kasungu and Lilongwe Districts of Malawi in 2019. Malawi Medical Journal. 2020;32(4):184-91.
62. Mbachu CN, Ebenebe JC, Okpara HC, Chukwuka JO, Mbachu II, Elo-Ilo JC, Ndukwu CI, Egbuonu I. Hepatitis e prevalence, knowledge, and practice of preventive measures among secondary school adolescents in rural Nigeria: a cross-sectional study. BMC Public Health. 2021 Dec;21:1-8.
63. Semá Baltazar C, Horth R, Boothe M, Sathane I, Young P, Chitsondzo Langa D, Condula M, Ricardo H, Dengo Baloi L, Cummings B, Schaad N. High prevalence of HIV, HBsAg and anti-HCV positivity among people who injected drugs: results of the first bio-behavioral survey using respondent-driven sampling in two urban areas in Mozambique. BMC infectious diseases. 2019 Dec;19:1-3.
64. Adoueni VK, Azoh AJ, Kouame E, Meless DG, Sibailly P, Derbe AK, Dzade KB, Koffi S, Kouakou T, Arra LV, Ouattara Y. Prevalence and correlates of hypothyroidism in pregnancy: a cross-sectional study at Bouget General Hospital, Ivory Coast. Pan African Medical Journal. 2022 Jan 13;41(1).
65. Gbeasor-Komlanvi FA, Zida-Compaore WI, Sadio AJ, Tchankoni MK, Kadangha BM, Salou M, Dagnra AC, Ekouevi DK. HIV testing uptake and prevalence among hospitalized older adults in Togo: A cross-sectional study. Plos one. 2021 Feb 2;16(2):e0246151.

100. Jonas A, Patel SV, Katuta F, Maher AD, Banda KM, Gerndt K, Pietersen I, de Prata NM, Mutenda N, Nakanyala T, Kisting E. HIV prevalence, risk factors for infection, and uptake of prevention, testing, and treatment among female sex workers in Namibia. Journal of epidemiology and global health. 2020 Dec;10(4):351.
101. Ngobeni R, Gilchrist C, Samie A. Prevalence and Distribution of Cryptosporidium spp. and Giardia lamblia in Rural and Urban Communities of South Africa. Turkiye parazitolojii dergisi. 2022 Mar 1;46(1):14-9.
102. Diriba K, Churiso G. The prevalence of Mycobacterium tuberculosis using Gene Xpert among tuberculosis suspected patients in Gedeo Zone, Southern Ethiopia. European Journal of Medical Research. 2022 Dec;27(1):1-8.
103. Shayo G, Makundi I, Luzzatto L. The prevalence of human immunodeficiency and of hepatitis B viral infections is not increased in patients with sickle cell disease in Tanzania. BMC Infectious Diseases. 2021 Dec;21:1-6.
104. Berhe T, Gebreyesus H, Teklay H. Prevalence and determinants of stillbirth among women attended deliveries in Aksum General Hospital: a facility based cross-sectional study. BMC research notes. 2019 Dec;12:1-6.
105. Mcharo RD, Kisinda A, Njovu L, Mcharo M, Mbwilo F, Mihale G, Komba B, Andrew E, Mayaud P, Kroidl A, Ivanova O. Prevalence of and risk factors associated with HIV, Herpes Simplex Virus-type 2, Chlamydia trachomatis and Neisseria gonorrhoeae infections among 18–24 year old students attending Higher Learning Institutions in Mbeya-Tanzania. Plos one. 2022 May 26;17(5):e0266596.
106. Chekesa B, Gumi B, Chanyalew M, Zewude A, Ameni G. Prevalence of latent tuberculosis infection and associated risk factors in prison in East Wollega Zone of western Ethiopia. PLoS One. 2020 May 19;15(5):e0233314.
107. Nanyonjo G, Asiki G, Ssetaala A, Nakaweesa T, Wambuzi M, Nanvubya A, Mpendo J, Okech B, Kitandwe PK, Nielsen L, Nalutaaya A. Prevalence and correlates of HIV infection among adolescents and young people living in fishing populations along Lake Victoria Fishing Communities in Uganda. Pan African Medical Journal. 2020 Nov 2;37(1).
108. Ntonifor NH, Tamufor AS, Abongwa LE. Prevalence of intestinal parasites and associated risk factors in HIV positive and negative patients in Northwest Region, Cameroon. Scientific Reports. 2022 Oct 6;12(1):16747.
109. Abaka-Yawson A, Sosu SQ, Kwadzokpui PK, Afari S, Adusei S, Arko-Mensah J. Prevalence and determinants of intestinal parasitic infections among pregnant women receiving antenatal care in Kasoa Polyclinic, Ghana. Journal of Environmental and Public Health. 2020 Sep 8;2020.
110. Fernández-Luis, S., Fuente-Soro, L., Nhampossa, T., Lopez-Varela, E., Augusto, O., Nhacolo, A., Vazquez, O., Saura-Lázaro, A., Guambe, H., Tibana, K. and Ngeno, B., 2022. Prompt HIV diagnosis and antiretroviral treatment in postpartum women is crucial for prevention of mother to child transmission during breastfeeding: Survey results in a high HIV prevalence community in southern Mozambique after the implementation of Option B+. *PloS one*, *17*(8), p.e0269835.
111. Hailu GG, Ayele ET. Assessment of the prevalence of intestinal parasitic infections and associated habit and culture-related risk factors among primary schoolchildren in Debre Berhan town, Northeast Ethiopia. BMC public health. 2021 Dec;21(1):1-2.
112. Alula GA, Munshea A, Nibret E. Prevalence of intestinal parasitic infections and associated risk factors among pregnant women attending prenatal care in the Northwestern Ethiopia. BioMed Research International. 2021 Dec 23;2021.
113. Ismail A, Darling AM, Mosha D, Fawzi W, Sudfeld C, Sando MM, Abdallah Noor R, Charles J, Vuai S. Prevalence and risk factors associated with malnutrition among adolescents in rural Tanzania. Tropical Medicine & International Health. 2020 Jan;25(1):89-100.
114. Abdo RA, Halil HM, Kebede BA, Anshebo AA, Gejo NG. Prevalence and contributing factors of birth asphyxia among the neonates delivered at Nigist Eleni Mohammed memorial teaching hospital, Southern Ethiopia: a cross-sectional study. BMC pregnancy and childbirth. 2019 Dec;19:1-7.
115. Ferrao J, Earland D, Novela A, Mendes R, Ballat M, Tungadza A, Searle K. Modelling sociodemographic factors that affect malaria prevalence in Sussundenga, Mozambique: a cross-sectional study. F1000Research. 2022;11.

116. Thistle P, Parpia R, Pain D, Lee H, Manasa J, Schnipper LE. Prevalence and subtype distribution of high-risk human papillomavirus among women presenting for cervical cancer screening at Karanda Mission Hospital. JCO Global Oncology. 2020 Aug;6:1276-81.
117. Mangusho C, Mwebesa E, Izudi J, Aleni M, Dricile R, Ayiasi RM, Legason ID. High prevalence of malaria in pregnancy among women attending antenatal care at a large referral hospital in northwestern Uganda: A cross-sectional study. Plos one. 2023 Apr 5;18(4):e0283755.
118. Korzeniewski K, Bylicka-Szczepanowska E, Lass A. Prevalence of asymptomatic malaria infections in seemingly healthy children, the rural Dzanga Sangha region, Central African Republic. International Journal of Environmental Research and Public Health. 2021 Jan;18(2):814.
119. Sacolo-Gwebu H, Chimbari M, Kalinda C. Prevalence and risk factors of schistosomiasis and soil-transmitted helminthiases among preschool aged children (1–5 years) in rural KwaZulu-Natal, South Africa: a cross-sectional study. Infectious diseases of poverty. 2019 Dec;8:1-2.
120. Melkie G, Azage M, Gedamu G. Prevalence and associated factors of active trachoma among children aged 1-9 years old in mass drug administration graduated and non-graduated districts in Northwest Amhara region, Ethiopia: A comparative cross-sectional study. Plos one. 2020 Dec 15;15(12):e0243863.
121. DeWyer A, Scheel A, Webel AR, Longenecker CT, Kamarembo J, Aliku T, Engel ME, Bowen AC, Bwanga F, Hovis I, Chang A. Prevalence of group A β-hemolytic streptococcal throat carriage and prospective pilot surveillance of streptococcal sore throat in Ugandan school children. International Journal of Infectious Diseases. 2020 Apr 1;93:245-51.
122. Abebaw A, Aschale Y, Kebede T, Hailu A. The prevalence of symptomatic and asymptomatic malaria and its associated factors in Debre Elias district communities, Northwest Ethiopia. Malaria Journal. 2022 Dec;21(1):1-0.
123. Cowley G, Milne G, da Silva ET, Nakutum J, Rodrigues A, Vasileva H, Mabey D, Versteeg B, Last A. Prevalence of and risk factors for curable sexually transmitted infections on Bubaque Island, Guinea Bissau. Sexually transmitted infections. 2021 Feb 1;97(1):51-5.
124. Mabunda N, Vieira L, Chelene I, Maueia C, Zicai AF, Duajá A, Chale F, Chambal L, Vubil A, Augusto O. Prevalence of hepatitis B virus and immunity status among healthcare workers in Beira City, Mozambique. Plos one. 2022 Oct 14;17(10):e0276283.
125. Girma T, Gezimu W, Demeke A. Prevalence, causes, and factors associated with obstructed labour among mothers who gave birth at public health facilities in Mojo Town, Central Ethiopia, 2019: A cross-sectional study. PLoS One. 2022 Sep 22;17(9):e0275170.
126. Abose S, Nuramo A, Brehane M, Lemma L, Ahemed R, Gebrehiwot H. The prevalence and associated factors of birth asphyxia among neonates delivered in Public Hospitals, Northern Ethiopia. African Health Sciences. 2022 Aug 1;22(2):518-25.
127. Shimelash A, Alemayehu M, Dagne H, Mihiretie G, Lamore Y, Tegegne E, Kumlachew L. Prevalence of active trachoma and associated factors among school age children in Debre Tabor Town, Northwest Ethiopia, 2019: a community based cross-sectional study. Italian Journal of Pediatrics. 2022 Dec;48(1):1-9.
128. Ribado Meñe G, Dejon Agobé JC, Momo Besahà JC, Abaga Ondo Ndoho F, Abdulla S, Adegnika AA. Prevalence, intensity and associated risk factors of soil-transmitted helminth infections among individuals living in Bata district, Equatorial Guinea. PLOS Neglected Tropical Diseases. 2023 May 17;17(5):e0011345.
129. Limenih A, Gelaye W, Alemu G. Prevalence of Malaria and Associated Factors among Delivering Mothers in Northwest Ethiopia. BioMed Research International. 2021 Dec 7;2021.
130. Tsegaye AT, Ayele A, Birhanu S. Prevalence and associated factors of malaria in children under the age of five years in Wogera district, northwest Ethiopia: A cross-sectional study. Plos one. 2021 Oct 11;16(10):e0257944.
131. Mlugu EM, Minzi O, Kamuhabwa AA, Aklillu E. Prevalence and correlates of asymptomatic malaria and anemia on first antenatal care visit among pregnant women in Southeast, Tanzania. International journal of environmental research and public health. 2020 May;17(9):3123.
132. Beavogui AH, Cherif MS, Camara BS, Delamou A, Kolie D, Cissé A, Camara D, Sow A, Camara G, Yattara M, Goumou N. Prevalence of parasitic infections in children of Boke, Guinea. The Journal of Parasitology. 2021 Sep 1;107(5):783-9.
133. Kayuni SA, O'Ferrall AM, Baxter H, Hesketh J, Mainga B, Lally D, Al-Harbi MH, LaCourse EJ, Juziwelo L, Musaya J, Makaula P. An outbreak of intestinal schistosomiasis, alongside increasing urogenital

schistosomiasis prevalence, in primary school children on the shoreline of Lake Malawi, Mangochi District, Malawi. Infectious Diseases of Poverty. 2020 Dec;9:1-0.
134. Kojom Foko LP, Nolla NP, Nyabeyeu Nyabeyeu H, Tonga C, Lehman LG. Prevalence, patterns, and determinants of malaria and malnutrition in Douala, Cameroon: a cross-sectional community-based study. BioMed Research International. 2021 Jul 12;2021:1-2.
135. Yohannes M, Abebe Z, Boelee E. Prevalence and environmental determinants of cutaneous leishmaniasis in rural communities in Tigray, northern Ethiopia. PLoS neglected tropical diseases. 2019 Sep 26;13(9):e0007722.
136. Njoya, H.F., Awolu, M.M., Christopher, T.B., Duclerc, J.F., Ateudjieu, J., Wirsiy, F.S., Atuhaire, C. and Cumber, S.N., 2021. Prevalence and awareness of mode of transmission of typhoid fever in patients diagnosed with Salmonella typhi and paratyphi infections at the Saint Elisabeth General Hospital Shisong, Bui Division, Cameroon. *Pan African Medical Journal*, *40*(1).
137. Abebaw A, Alemu G, Ayehu A. Prevalence of intestinal parasites and associated factors among children from child centres in Bahir Dar city, northwest Ethiopia. Tropical doctor. 2020 Jul;50(3):194-8.
138. Aschale A, Adane M, Getachew M, Faris K, Gebretsadik D, Sisay T, Dewau R, Chanie MG, Muche A, Zerga AA, Lingerew M. Water, sanitation, and hygiene conditions and prevalence of intestinal parasitosis among primary school children in Dessie City, Ethiopia. Plos one. 2021 Feb 3;16(2):e0245463.
139. Chilongola JO, Sabuni EJ, Kapyolo EP. Prevalence of plasmodium, leptospira and rickettsia species in Northern Tanzania: a community based survey. African Health Sciences. 2020 Apr 20;20(1):199-207.
140. Mohamed AK, Elhassan NM, Awhag ZA, Ali FS, Ali ET, Mhmoud NA, Siddig EE, Hassan R, Ahmed ES, Fattahi A, Ahmed A. Prevalence of Helicobacter pylori among Sudanese patients diagnosed with colon polyps and colon cancer using immunohistochemistry technique. BMC Research Notes. 2020 Dec;13:1-6.
141. Hajare ST, Gobena RK, Chauhan NM, Eriso F. Prevalence of intestinal parasite infections and their associated factors among food handlers working in selected catering establishments from Bule Hora, Ethiopia. BioMed Research International. 2021 Aug 19;2021.
142. Mazigo HD, Mwingira UJ, Zinga MM, Uisso C, Kazyoba PE, Kinung'hi SM, Mutapi F. Urogenital schistosomiasis among pre-school and school aged children in four districts of north western Tanzania after 15 years of mass drug administration: geographical prevalence, risk factors and performance of haematuria reagent strips. PLOS Neglected Tropical Diseases. 2022 Oct 12;16(10):e0010834.
143. Bassa FK, Eze IC, Assaré RK, Essé C, Koné S, Acka F, Laubhouet-Koffi V, Kouassi D, Bonfoh B, Utzinger J, N'Goran EK. Prevalence of Schistosoma mono-and co-infections with multiple common parasites and associated risk factors and morbidity profile among adults in the Taabo health and demographic surveillance system, South-Central Côte d'Ivoire. Infectious diseases of poverty. 2022 Dec;11:1-2.
144. Mwebaza S, Senyonga B, Atuhairwe C, Taremwa IM. Prevalence and associated factors of intestinal parasitic infections among HIV clients attending Masaka Regional Referral Hospital, Uganda. The Pan African Medical Journal. 2022 Nov 4;43(122).
145. Awol RN, Reda DY, Gidebo DD. Prevalence of Salmonella enterica serovar Typhi infection, its associated factors and antimicrobial susceptibility patterns among febrile patients at Adare general hospital, Hawassa, southern Ethiopia. BMC infectious diseases. 2021 Dec;21:1-9.
146. Adjobimey M, Ade S, Wachinou P, Esse M, Yaha L, Bekou W, Campbell JR, Toundoh N, Adjibode O, Attikpa G, Agodokpessi G. Prevalence, acceptability, and cost of routine screening for pulmonary tuberculosis among pregnant women in Cotonou, Benin. PLoS One. 2022 Feb 22;17(2):e0264206.
147. Bule YP, Silva J, Carrilho C, Campos C, Sousa H, Tavares A, Medeiros R. Human papillomavirus prevalence and distribution in self-collected samples from female university students in Maputo. International Journal of Gynecology & Obstetrics. 2020 May;149(2):237-46.
148. Bramania PK, Ruggajo P, Bramania R, Mahmoud M, Furia FF. Prevalence of malnutrition inflammation complex syndrome among patients on maintenance haemodialysis at Muhimbili National Hospital in Tanzania: a cross-sectional study. BMC nephrology. 2020 Dec;21(1):1-1.
149. Moglad EH, Ahmed DA, Awad AL-Kareem SM, Elgoraish AG, Ali HT, Altayb HN. Prevalence of human immunodeficiency virus among pulmonary tuberculosis patients: A cross-sectional study. Microbiology and Immunology. 2020 Dec;64(12):810-4.

150. Oppong M, Lamptey H, Kyei-Baafour E, Aculley B, Ofori EA, Tornyigah B, Kweku M, Ofori MF. Prevalence of sickle cell disorders and malaria infection in children aged 1–12 years in the Volta Region, Ghana: a community-based study. Malaria Journal. 2020 Dec;19(1):1-1.
151. Mushi V, Zacharia A, Shao M, Mubi M, Tarimo D. Prevalence and risk factors of urogenital schistosomiasis among under-fives in Mtama District in the Lindi region of Tanzania. PLoS Neglected Tropical Diseases. 2022 Apr 20;16(4):e0010381.
152. Daka D, Hailemeskel G, Fenta DA. Prevalence of Hepatitis B Virus infection and associated factors among female sex workers using respondent-driven sampling in Hawassa City, Southern Ethiopia. BMC microbiology. 2022 Jan 31;22(1):37.
153. Ayele A, Abera D, Hailu M, Birhanu M, Desta K. Prevalence and associated risk factors for Hepatitis B and C viruses among refugees in Gambella, Ethiopia. BMC public health. 2020 Dec;20(1):1-0.
154. Badiane AS, Ndiaye T, Thiaw AB, Binta DA, Diallo MA, Seck MC, Diongue K, Garba MN, Ndiaye M, Ndiaye D. High prevalence of asymptomatic Plasmodium infection in Bandafassi, South-East Senegal. Malaria Journal. 2021 May 12;20(1):218.
155. Oseni Okolo ML, Omatola CA. Hepatitis B and syphilis prevalence and risk factors of transmission among febrile patients in a primary health facility in Kogi State, Nigeria. Journal of Immunoassay and Immunochemistry. 2022 Jan 2;43(1):1938607.
156. Getie M, Abebe W, Tessema B. Prevalence of enteric bacteria and their antimicrobial susceptibility patterns among food handlers in Gondar town, Northwest Ethiopia. Antimicrobial Resistance & Infection Control. 2019 Dec;8(1):1-6.
157. Tadesse S, Mulu W, Genet C, Kibret M, Belete MA. Emergence of high prevalence of extended-spectrum beta-lactamase and carbapenemase-producing Enterobacteriaceae species among patients in Northwestern Ethiopia Region. BioMed Research International. 2022 Feb 4;2022.
158. Zeynudin A, Degefa T, Tesfaye M, Suleman S, Yesuf EA, Hajikelil Z, Ali S, Azam K, Husen A, Yasin J, Wieser A. Prevalence and intensity of soil-transmitted helminth infections and associated risk factors among household heads living in the peri-urban areas of Jimma town, Oromia, Ethiopia: A community-based cross-sectional study. Plos one. 2022 Sep 15;17(9):e0274702.
159. Ayele BH, Geleto A, Ayana DA, Redi M. Prevalence of feco-oral transmitted protozoan infections and associated factors among university students in Ethiopia: a cross-sectional study. BMC infectious diseases. 2019 Dec;19:1-8.
160. Mohamadou M, Essama SR, Ngonde Essome MC, Akwah L, Nadeem N, Gonsu Kamga H, Sattar S, Javed S. High prevalence of Panton-Valentine leukocidin positive, multidrug resistant, Methicillin-resistant Staphylococcus aureus strains circulating among clinical setups in Adamawa and Far North regions of Cameroon. Plos one. 2022 Jul 8;17(7):e0265118.
161. Ameha Z, Tadesse S, Assefa A, Tessema B. Prevalence and associated factors of Hepatitis C virus and human immunodeficiency virus infections among voluntary counseling and testing clients attending private health facilities in Bahir Dar city, North West Ethiopia 2014. BMC Research Notes. 2019 Dec;12(1):1-6.
162. Mooney JP, DonVito SM, Jahateh M, Bittaye H, Bottomley C, D’Alessandro U, Riley EM. Dry season prevalence of Plasmodium falciparum in asymptomatic gambian children, with a comparative evaluation of diagnostic methods. Malaria journal. 2022 Jun 7;21(1):171.
163. Ajayi AI, Ahinkorah BO, Seidu AA, Adeniyi OV. Prevalence and correlates of induced abortion: results of a facility-based cross-sectional survey of parturient women living with HIV in South Africa. Reproductive Health. 2022 Dec 5;19(1):220.
164. Tibebu EA, Desta KW, Ashagre FM, Jemberu AA. Prevalence of birth injuries and associated factors among newborns delivered in public hospitals Addis Ababa, Ethiopia, 2021. Crossectional study. Plos one. 2023 Jan 30;18(1):e0281066.
165. Nyumayo S, Konje E, Kidenya B, Kapesa A, Hingi M, Wango N, Ngimbwa J, Alphonce V, Basinda N. Prevalence of HIV and associated risk factors among street-connected children in Mwanza city. PLoS One. 2022 Nov 8;17(11):e0271042.
166. Adane MM, Alene GD, Mereta ST, Wanyonyi KL. Prevalence and risk factors of acute lower respiratory infection among children living in biomass fuel using households: a community-based cross-sectional study in Northwest Ethiopia. BMC Public Health. 2020 Dec;20:1-3.

167. Chilanga E, Collin-Vezina D, MacIntosh H, Mitchell C, Cherney K. Prevalence and determinants of malaria infection among children of local farmers in Central Malawi. Malaria Journal. 2020 Dec;19(1):1-0.
168. Folarin OF, Kuti BP, Oyelami AO. Prevalence, density and predictors of malaria parasitaemia among ill young Nigerian infants. Pan African Medical Journal. 2021 Sep 9;40(1).
169. Ukke GG, Diriba K. Prevalence and factors associated with neonatal hypothermia on admission to neonatal intensive care units in Southwest Ethiopia–a cross-sectional study. PloS one. 2019 Jun 6;14(6):e0218020.
170. Itaka MB, Omole OB. Prevalence and factors associated with malnutrition among under 5-year-old children hospitalised in three public hospitals in South Africa. African Journal of Primary Health Care and Family Medicine. 2020 Jan 1;12(1):1-7.
171. Alemu Y, Degefa T, Bajiro M, Teshome G. Prevalence and intensity of soil-transmitted helminths infection among individuals in model and non-model households, South West Ethiopia: A comparative cross-sectional community based study. Plos one. 2022 Oct 17;17(10):e0276137.
172. Rahantamalala A, Rakotoarison RL, Rakotomalala E, Rakotondrazaka M, Kiernan J, Castle PM, Hakami L, Choi K, Rafalimanantsoa AS, Harimanana A, Wright P. Prevalence and factors associated with human Taenia solium taeniosis and cysticercosis in twelve remote villages of Ranomafana rainforest, Madagascar. PLoS Neglected Tropical Diseases. 2022 Apr 11;16(4):e0010265.
173. Kawambwa RH, Majigo MV, Mohamed AA, Matee MI. High prevalence of human immunodeficiency virus, hepatitis B and C viral infections among people who inject drugs: a potential stumbling block in the control of HIV and viral hepatitis in Tanzania. BMC Public Health. 2020 Dec;20:1-7.
174. Ijoma UN, Meka IA, Omotowo B, Nwagha TU, Obienu O, Onodugo OD, Onyekonwu CL, Okoli EV, Ndu AC, Ugwu EO. Sero-prevalence of Hepatitis B virus infection: A cross-sectional study of a large population of health care workers in Nigeria. Nigerian Journal of Clinical Practice. 2021 Jan 1;24(1):38-44.
175. Duguma T, Tekalign E, Muleta D, Simieneh A. Malaria prevalence and risk factors among patients visiting Mizan Tepi University Teaching Hospital, Southwest Ethiopia. Plos one. 2022 Jul 28;17(7):e0271771.
176. Woday A, Mohammed A, Gebre A, Urmale K. Prevalence and associated factors of malaria among febrile children in Afar region, Ethiopia: a health facility based study. Ethiopian journal of health sciences. 2019;29(5).
177. Bancha B, Kinfe AA, Chanko KP, Workie SB, Tadese T. Prevalence of hepatitis B viruses and associated factors among pregnant women attending antenatal clinics in public hospitals of Wolaita Zone, South Ethiopia. PloS one. 2020 May 7;15(5):e0232653.
178. Amsalu T, Genet C, Adem Siraj Y. Salmonella Typhi and S almonella Paratyphi prevalence, antimicrobial susceptibility profile and factors associated with enteric fever infection in Bahir Dar, Ethiopia. Scientific Reports. 2021 Apr 1;11(1):7359.
179. Adeniji EO, Kuti BP, Elusiyan JB. Prevalence, risk factors, and outcome of hospitalization of neonatal hyperglycemia at a Nigerian health facility. Nigerian Journal of Clinical Practice. 2020 Jan 1;23(1):71-8.
180. Govender K, Beckett S, Reddy T, Cowden RG, Cawood C, Khanyile D, Kharsany AB, George G, Puren A. Association of HIV Intervention uptake with HIV prevalence in adolescent girls and young women in South Africa. JAMA Network Open. 2022 Apr 1;5(4):e228640-.
181. WoldeKidan E, Daka D, Legesse D, Laelago T, Betebo B. Prevalence of active trachoma and associated factors among children aged 1 to 9 years in rural communities of Lemo district, southern Ethiopia: community based cross sectional study. BMC infectious diseases. 2019 Dec;19:1-8.
182. Conan N, Coulborn RM, Simons E, Mapfumo A, Apollo T, Garone DB, Casas EC, Puren AJ, Chihana ML, Maman D. Successes and gaps in the HIV cascade of care of a high HIV prevalence setting in Zimbabwe: a population-based survey. Journal of the International AIDS Society. 2020 Sep;23(9):e25613.
183. Megersa T, Dango S, Kumsa K, Lemma K, Lencha B. Prevalence of high-risk human papillomavirus infections and associated factors among women living with HIV in Shashemene town public health facilities, Southern Ethiopia. BMC Women's Health. 2023 Mar 23;23(1):125.
184. Vueba AN, Faria CP, Almendra R, Santana P, Sousa MD. Serological prevalence of toxoplasmosis in pregnant women in Luanda (Angola): Geospatial distribution and its association with socio-demographic and clinical-obstetric determinants. Plos one. 2020 Nov 6;15(11):e0241908.
185. Tchankoni MK, Bitty-Anderson AM, Sadio AJ, Gbeasor-Komlanvi FA, Ferré VM, Zida-Compaore WI, Dorkenoo AM, Saka B, Dagnra AC, Charpentier C, Ekouevi DK. Prevalence and factors associated with

trichomonas vaginalis infection among female sex workers in Togo, 2017. BMC Infectious Diseases. 2021 Dec;21(1):1-7.

186. Karau PB, Kirna B, Amayo E, Joshi M, Ngare S, Muriira G. The prevalence of vitamin D deficiency among patients with type 2 diabetes seen at a referral hospital in Kenya. Pan African Medical Journal. 2019 Sep 17;34(1).
187. Tola MA, Abera NA, Gebeyehu YM, Dinku SF, Tullu KD. High prevalence of extended-spectrum beta-lactamase-producing Escherichia coli and Klebsiella pneumoniae fecal carriage among children under five years in Addis Ababa, Ethiopia. PloS one. 2021 Oct 1;16(10):e0258117.
188. Kara WS, Chikomele J, Mzigaba MM, Mao J, Mghanga FP. Anaemia in pregnancy in Southern Tanzania: Prevalence and associated risk factors. African Journal of Reproductive Health. 2020 Dec 3;24(3):154-60.
189. Abera W, Gintamo B, Shitemaw T, Mekuria ZN, Gizaw Z. Prevalence of intestinal parasites and associated factors among food handlers in food establishments in the Lideta subcity of Addis Ababa, Ethiopia: an institution-based, cross-sectional study. BMJ open. 2022 Jul 1;12(7):e061688.
190. Terefe N, Nigussie A, Tadele A. Prevalence of obstetric danger signs during pregnancy and associated factors among mothers in Shashemene Rural District, South Ethiopia. Journal of Pregnancy. 2020 Sep 26;2020.
191. Asfaw MA, Gezmu T, Wegayehu T, Bekele A, Hailemariam Z, Masresha N, Gebre T. Soil-transmitted helminth infections among pre-school aged children in Gamo Gofa zone, Southern Ethiopia: Prevalence, intensity and intervention status. Plos one. 2020 Dec 15;15(12):e0243946.
192. Rumisha SF, Shayo EH, Mboera LE. Spatio-temporal prevalence of malaria and anaemia in relation to agro-ecosystems in Mvomero district, Tanzania. Malaria Journal. 2019 Dec;18(1):1-4.
193. Hope D, Businge S, Kyoyagala S, Bazira J. Prevalence of anti-leptospiral IgM and detection of pathogenic Leptospira species DNA in neonates presenting with clinical sepsis in Southwestern Uganda. European Journal of Medical Research. 2022 Dec;27(1):1-8.
194. Salgado C, Ayodo G, Macklin MD, Gould MP, Nallandhighal S, Odhiambo EO, Obala A, O’Meara WP, John CC, Tran TM. The prevalence and density of asymptomatic Plasmodium falciparum infections among children and adults in three communities of western Kenya. Malaria Journal. 2021 Dec;20:1-1.
195. Kebede D, Admas A, Mekonnen D. Prevalence and antibiotics susceptibility profiles of Streptococcus pyogenes among pediatric patients with acute pharyngitis at Felege Hiwot Comprehensive Specialized Hospital, Northwest Ethiopia. BMC microbiology. 2021 Dec;21(1):1-0.
196. Masaku J, Njomo DW, Njoka A, Okoyo C, Mutungi FM, Njenga SM. Soil-transmitted helminths and schistosomiasis among pre-school age children in a rural setting of Busia County, Western Kenya: a cross-sectional study of prevalence, and associated exposures. BMC public health. 2020 Dec;20(1):1-1.
197. Sigei L, Nyaga EM, Milimo B. Prevalence and immediate outcomes of low birth weight neonates born of pre-eclamptic women at Moi Teaching and Referral Hospital, Kenya. The Pan African Medical Journal. 2023;44.
198. Eyayu T, Kiros T, Workineh L, Sema M, Damtie S, Hailemichael W, Dejen E, Tiruneh T. Prevalence of intestinal parasitic infections and associated factors among patients attending at Sanja Primary Hospital, Northwest Ethiopia: An institutional-based cross-sectional study. PLoS One. 2021 Feb 16;16(2):e0247075.
199. Mbuya AW, Mboya IB, Semvua HH, Mamuya SH, Msuya SE. Prevalence and factors associated with tuberculosis among the mining communities in Mererani, Tanzania. Plos one. 2023 Mar 15;18(3):e0280396.
200. Msollo SS, Martin HD, Mwanri AW, Petrucka P. Prevalence of hyperglycemia in pregnancy and influence of body fat on development of hyperglycemia in pregnancy among pregnant women in urban areas of Arusha region, Tanzania. BMC pregnancy and childbirth. 2019 Dec;19(1):1-9.
201. Mnkugwe RH, Minzi OS, Kinung'hi SM, Kamuhabwa AA, Aklillu E. Prevalence and correlates of intestinal schistosomiasis infection among school-aged children in North-Western Tanzania. PloS one. 2020 Feb 5;15(2):e0228770.
202. Misikir SW, Wobie M, Tariku MK, Bante SA. Prevalence of hookworm infection and associated factors among pregnant women attending antenatal care at governmental health centers in DEMBECHA district, north West Ethiopia, 2017. BMC Pregnancy and Childbirth. 2020 Dec;20:1-8.
203. Chihana ML, Conan N, Ellman T, Poulet E, Garone DB, Ortuno R, Wanjala S, Masiku C, Etard JF, Davies MA, Maman DJ. The HIV cascade of care among serodiscordant couples in four high HIV prevalence settings in sub-Saharan Africa. South African Medical Journal. 2021 Aug 1;111(8):768-76.

204. Mang'ara RJ, Ngasala B, John W. Prevalence of Schistosoma mansoni infection among fishermen in Busega district, Tanzania. Plos one. 2022 Nov 28;17(11):e0276395.
205. Nyamu GW, Kihara JH, Oyugi EO, Omballa V, El-Busaidy H, Jeza VT. Prevalence and risk factors associated with asymptomatic Plasmodium falciparum infection and anemia among pregnant women at the first antenatal care visit: a hospital based cross-sectional study in Kwale County, Kenya. PloS one. 2020 Oct 8;15(10):e0239578.
206. Donkoh ET, Asmah RH, Agyemang-Yeboah F, Dabo EO, Wiredu EK. Prevalence and distribution of vaccine-preventable genital human Papillomavirus (Hpv) genotypes in ghanaian women presenting for screening. Cancer Control. 2022 Apr 20;29:10732748221094721.
207. Muche AA, Olayemi OO, Gete YK. Prevalence of gestational diabetes mellitus and associated factors among women attending antenatal care at Gondar town public health facilities, Northwest Ethiopia. BMC pregnancy and childbirth. 2019 Dec;19:1-3.
208. Mazigo HD, Uisso C, Kazyoba P, Nshala A, Mwingira UJ. Prevalence, infection intensity and geographical distribution of schistosomiasis among pre-school and school aged children in villages surrounding Lake Nyasa, Tanzania. Scientific Reports. 2021 Jan 11;11(1):295.
209. Ajepe AA, Okunade KS, Sekumade AI, Daramola ES, Beke MO, Ijasan O, Olowoselu OF, Afolabi BB. Prevalence and foetomaternal effects of iron deficiency anaemia among pregnant women in Lagos, Nigeria. PLoS One. 2020 Jan 23;15(1):e0227965.
210. Chiesa A, Ochola E, Oreni L, Vassalini P, Rizzardini G, Galli M. Hepatitis B and HIV coinfection in Northern Uganda: Is a decline in HBV prevalence on the horizon?. PloS one. 2020 Nov 18;15(11):e0242278.
211. Woldegiorgis AE, Erku W, Medhin G, Berhe N, Legesse M. Community-based sero-prevalence of hepatitis B and C infections in South Omo Zone, Southern Ethiopia. PloS one. 2019 Dec 30;14(12):e0226890.
212. Djuikoue IC, Tambo E, Tazemda G, Njajou O, Makoudjou D, Sokeng V, Wandji M, Tomi C, Nanfack A, Dayomo A, Lacmago S. Evaluation of inpatients Clostridium difficile prevalence and risk factors in Cameroon. Infectious Diseases of Poverty. 2020 Dec;9:1-7.
213. Ojerinde OA, Ojo SK, Udewena UL, Oladeji SJ. A cross-sectional study on the prevalence of HIV and hepatitis B virus co-infection among students of a tertiary institution in Ekiti State, Southwest Nigeria. The Pan African Medical Journal. 2023;44.
214. Kaba D, Bangoura MA, Sylla MM, Sako FB, Diallo MS, Diallo I, Kolié OO, Keita AS, Diané BF, Keita F, Diakité M. Prevalence and factors associated with hepatitis B in a cohort of HIV-infected children in the Pediatric Department at Donka National Hospital, Guinea. Pan African Medical Journal. 2019 Dec 6;34(1).
215. Tesfa H, Jara D, Woyiraw W, Bogale EK, Asrat B. Prevalence of undernourishment and associated factors among adults with major depressive disorder at two public hospitals in Northwest Ethiopia: a cross-sectional study. BMJ open. 2022 Nov 1;12(11):e065108.
216. Birjandi MM, Oroei M. The prevalence of positive rapid diagnostic test of hepatitis C virus infection in Ghana. Pan African Medical Journal. 2020 Aug 21;36(1).
217. Machano MM, Joho AA. Prevalence and risk factors associated with severe pre-eclampsia among postpartum women in Zanzibar: a cross-sectional study. BMC Public Health. 2020 Dec;20:1-0.
218. September J, Geffen L, Manning K, Naicker P, Faro C, Mendelson M, Wasserman S. Colonisation with pathogenic drug-resistant bacteria and Clostridioides difficile among residents of residential care facilities in Cape Town, South Africa: a cross-sectional prevalence study. Antimicrobial Resistance & Infection Control. 2019 Dec;8(1):1-8.
219. Tekalign E, Bajiro M, Ayana M, Tiruneh A, Belay T. Prevalence and intensity of soil-transmitted helminth infection among rural community of southwest Ethiopia: a community-based study. BioMed Research International. 2019 Dec 14;2019.
220. Muluneh C, Hailu T, Alemu G. Prevalence and associated factors of soil-transmitted helminth infections among children living with and without open defecation practices in Northwest Ethiopia: a comparative cross-sectional study. The American journal of tropical medicine and hygiene. 2020 Jul;103(1):266.
221. Akala HM, Watson OJ, Mitei KK, Juma DW, Verity R, Ingasia LA, Opot BH, Okoth RO, Chemwor GC, Juma JA, Mwakio EW. Plasmodium interspecies interactions during a period of increasing prevalence of Plasmodium ovale in symptomatic individuals seeking treatment: an observational study. The Lancet Microbe. 2021 Apr 1;2(4):e141-50.

222. Sinshaw W, Kebede A, Bitew A, Tesfaye E, Tadesse M, Mehamed Z, Yenew B, Amare M, Dagne B, Diriba G, Alemu A. Prevalence of tuberculosis, multidrug resistant tuberculosis and associated risk factors among smear negative presumptive pulmonary tuberculosis patients in Addis Ababa, Ethiopia. BMC infectious diseases. 2019 Dec;19:1-5.
223. Kayambankadzanja RK, Schell CO, Namboya F, Phiri T, Banda-Katha G, Mndolo SK, Bauleni A, Castegren M, Baker T. The prevalence and outcomes of sepsis in adult patients in two hospitals in Malawi. The American Journal of Tropical Medicine and Hygiene. 2020 Apr;102(4):896.
224. Nang DW, Tukirinawe H, Okello M, Tayebwa B, Theophilus P, Sikakulya FK, Fajardo Y, Afodun AM, Kajabwangu R. Prevalence of high-risk human papillomavirus infection and associated factors among women of reproductive age attending a rural teaching hospital in western Uganda. BMC Women's Health. 2023 Dec;23(1):1-8.
225. Kejela T, Dekosa F. High prevalence of MRSA and VRSA among inpatients of Mettu Karl Referral Hospital, Southwest Ethiopia. Tropical Medicine & International Health. 2022 Aug;27(8):735-41.
226. Otuli Noël L, Nguma Jean-Didier B, Alongo Mike-Antoine M, Bosunga Gedeon K, Mukonkole Jean-Paulin M, Likwela Joris L, Okenge Jean-Pascal M. Prevalence of congenital malaria in Kisangani, a stable malaria transmission area in Democratic Republic of the Congo. Infectious Diseases in Obstetrics and Gynecology. 2020 Feb 25;2020.
227. Gruninger SK, Rasamoelina T, Rakotoarivelo RA, Razafindrakoto AR, Rasolojaona ZT, Rakotozafy RM, Soloniaina PR, Rakotozandrindrainy N, Rausche P, Doumbia CO, Jaeger A. Prevalence and risk distribution of schistosomiasis among adults in Madagascar: a cross-sectional study. Infectious Diseases of Poverty. 2023 Dec;12(1):1-0.
228. Ayensu J, Annan R, Lutterodt H, Edusei A, Peng LS. Prevalence of anaemia and low intake of dietary nutrients in pregnant women living in rural and urban areas in the Ashanti region of Ghana. Plos one. 2020 Jan 24;15(1):e0226026.
229. Kebede ZT, Yigezaw GS, Yilma TM, Delele TG. Prevalence of pregnancy-related complications and associated factors among reproductive-aged women in northwest Ethiopia: A community-based retrospective cross-sectional study. International Journal of Gynecology & Obstetrics. 2021 Jul;154(1):62-71.
230. Debash H, Bisetegn H, Ebrahim H, Feleke DG, Gedefie A, Tilahun M, Shibabaw A, Ebrahim E, Fiseha M, Abeje G. Prevalence and associated risk factors of malaria among febrile under-five children visiting health facilities in Ziquala district, Northeast Ethiopia: A multicenter cross-sectional study. PLoS One. 2022 Oct 27;17(10):e0276899.
231. Otu AA, Udoh UA, Ita OI, Hicks JP, Ukpeh I, Walley J. Prevalence of Zika and malaria in patients with fever in secondary healthcare facilities in south-eastern Nigeria. Tropical doctor. 2020 Jan;50(1):22-30.
232. Almaw A, Yimer M, Alemu M, Tegegne B. Prevalence of malaria and associated factors among symptomatic pregnant women attending antenatal care at three health centers in north-west Ethiopia. Plos one. 2022 Apr 7;17(4):e0266477.
233. Lendoye E, Ngoungou EB, Komba OM, Ollomo B, Bekale S, Yacka-Mouele L, Obounou BW, Ntyonga-Pono MP, Ngou-Milama E. Prevalence and factors associated to gestational diabetes mellitus among pregnant women in Libreville: a cross-sectional study. Pan African Medical Journal. 2022 Feb 15;41(1).
234. Mghanga F, Maduhu E, Nyawale H. Prevalence and associated factors of gestational diabetes mellitus among rural pregnant women in southern Tanzania. Ghana Medical Journal. 2020 Jun 30;54(2):82-7.
235. Ayele TB, Moyehodie YA. Prevalence of preterm birth and associated factors among mothers who gave birth in public hospitals of east Gojjam zone, Ethiopia. BMC Pregnancy and Childbirth. 2023 Mar 24;23(1):204.
236. Eneyew B, Sisay T, Gizeyatu A, Lingerew M, Keleb A, Malede A, Ademas A, Dagne M, Gebrehiwot M, Damtie Y, Tegegne TB. Prevalence and associated factors of acute respiratory infection among street sweepers and door-to-door waste collectors in Dessie City, Ethiopia: a comparative cross-sectional study. PloS one. 2021 May 14;16(5):e0251621.
237. Fatunla OA, Olatunya OS, Ogundare EO, Fatunla TO, Babatola AO, Adeniyi AT, Oyelami OA. Malaria prevention practices and malaria prevalence among children living in a rural community in Southwest Nigeria. The Journal of Infection in Developing Countries. 2022 Feb 28;16(02):352-61.
238. Akazong E, Tume C, Njouom R, Ayong L, Fondoh V, Kuiate JR. Knowledge, attitude and prevalence of hepatitis B virus among healthcare workers: a cross-sectional, hospital-based study in Bamenda Health District, NWR, Cameroon. BMJ open. 2020 Mar 1;10(3):e031075.

239. Mwaniki SW, Kaberia PM, Mugo PM, Palanee-Phillips T. HIV prevalence and associated risk factors among young tertiary student men who have sex with men (MSM) in Nairobi, Kenya: a respondent-driven sampling survey. AIDS Research and Therapy. 2023 Feb 6;20(1):7.
240. Bayih WA, Tezera TG, Alemu AY, Belay DM, Hailemeskel HS, Ayalew MY. Prevalence and determinants of asphyxia neonatorum among live births at Debre Tabor General Hospital, North Central Ethiopia: a cross-sectional study. African Health Sciences. 2021 Apr 16;21(1):385-96.
241. Ramatlho P, Grover S, Mathoma A, Tawe L, Matlhagela K, Ngoni K, Molebatsi K, Chilisa B, Zetola NM, Robertson ES, Paganotti GM. Human papillomavirus prevalence among unvaccinated young female college students in Botswana: A cross-sectional study. South African Medical Journal. 2022 May 1;112(5):335-40.
242. Belay A, Ashagrie M, Seyoum B, Alemu M, Tsegaye A. Prevalence of enteric pathogens, intestinal parasites and resistance profile of bacterial isolates among HIV infected and non-infected diarrheic patients in Dessie Town, Northeast Ethiopia. PLoS One. 2020 Dec 15;15(12):e0243479.
243. Desta ML, Saravanan M, Hilekiros H, Kahsay AG, Mohamed NF, Gezahegn AA, Lopes BS. HIV prevalence and risk factors in infants born to HIV positive mothers, measured by dried blood spot real-time PCR assay in Tigray, Northern Ethiopia. BMC pediatrics. 2019 Dec;19:1-8.
244. Scheibe A, Young K, Versfeld A, Spearman CW, Sonderup MW, Prabdial-Sing N, Puren A, Hausler H. Hepatitis B, hepatitis C and HIV prevalence and related sexual and substance use risk practices among key populations who access HIV prevention, treatment and related services in South Africa: findings from a seven-city cross-sectional survey (2017). BMC Infectious Diseases. 2020 Dec;20:1-5.
245. Oyedeji GJ, Adeyemo C, Dissou A, Abiodun T, Alli OA, Onaolapo OJ, Onaolapo AY, Adesiji Y, Olowe OA. Prevalence of multi-drug resistant tuberculosis among tuberculosis patients attending chest clinics in Osun-State, Nigeria. Current Pharmaceutical Biotechnology. 2020 Aug 1;21(10):939-47.
246. Yendewa GA, Lakoh S, Yendewa SA, Bangura K, Lawrence H, Patiño L, Jiba DF, Vandy AO, Murray MJ, Massaquoi SP, Deen GF. Prevalence of hepatitis B surface antigen and serological markers of other endemic infections in HIV-infected children, adolescents and pregnant women in Sierra Leone: A cross-sectional study. International Journal of Infectious Diseases. 2021 Jan 1;102:45-52.
247. Belete YA, Kassa TY, Baye MF. Prevalence of intestinal parasite infections and associated risk factors among patients of Jimma health center requested for stool examination, Jimma, Ethiopia. Plos one. 2021 Feb 22;16(2):e0247063.
248. Gitore WA, Ali MM, Yoseph A, Mangesha AE, Debiso AT. Prevalence of soil-transmitted helminthes and its association with water, sanitation, hygiene among schoolchildren and barriers for schools level prevention in technology villages of Hawassa University: Mixed design. Plos one. 2020 Sep 24;15(9):e0239557.
249. Endale A, Michlmayr D, Abegaz WE, Asebe G, Larrick JW, Medhin G, Legesse M. Community-based sero-prevalence of chikungunya and yellow fever in the South Omo Valley of Southern Ethiopia. PLoS Neglected Tropical Diseases. 2020 Sep 3;14(9):e0008549.
250. Yigezu H, Temam J, Bajiro M, Tesfaye Jule L, Nagaprasad N, Roy A, Saka A, Ramaswamy K. Factors Associated with the Prevalence of Hepatitis B among Volunteer Blood Donors at Jimma Blood Bank, South Ethiopia. Canadian Journal of Gastroenterology and Hepatology. 2022 May 23;2022.
251. Ajong AB, Kenfack B, Ali IM, Yakum MN, Telefo PB. Prevalence and correlates of low serum calcium in late pregnancy: A cross sectional study in the Nkongsamba Regional Hospital; Littoral Region of Cameroon. PloS one. 2019 Nov 7;14(11):e0224855.
252. Gadisa E, Jote K. Prevalence and factors associated with intestinal parasitic infection among under-five children in and around Haro Dumal Town, Bale Zone, Ethiopia. BMC pediatrics. 2019 Dec;19(1):1-8.
253. Ishungisa MA, Moen K, Leyna G, Makyao N, Ramadhan A, Lange T, Meyrowitsch DW, Mizinduko M, Likindikoki S, Leshabari M, Mmbaga EJ. HIV prevalence among men who have sex with men following the implementation of the HIV preventive guideline in Tanzania: respondent-driven sampling survey. BMJ open. 2020 Oct 1;10(10):e036460.
254. Wolde J, Haile D, Paulos K, Alemayehu M, Adeko AC, Ayza A. Prevalence of stillbirth and associated factors among deliveries attended in health facilities in Southern Ethiopia. Plos one. 2022 Dec 13;17(12):e0276220.
255. Omonijo AO, Omonijo A, Okoh HI, Ibrahim AO. Relationship between the usage of long-lasting insecticide-treated bed nets (LLITNs) and malaria prevalence among school-age children in Southwestern Nigeria. Journal of Environmental and Public Health. 2021 Mar 26;2021.

256. Nyitray AG, Masunaga K, Nyoni J, Ross MW. Prevalence of and factors associated with anal high-risk human papillomavirus in urban Tanzanian men who have sex with men, 2011-2012. International journal of STD & AIDS. 2022 Jun;33(7):672-9.
257. Mohamed N, Muse A, Wordofa M, Abera D, Mesfin A, Wolde M, Desta K, Tsegaye A, Taye B. Increased prevalence of cestode infection associated with history of deworming among primary school children in Ethiopia. The American Journal of Tropical Medicine and Hygiene. 2019 Sep;101(3):641.
258. Wemakor A. Prevalence and determinants of anaemia in pregnant women receiving antenatal care at a tertiary referral hospital in Northern Ghana. BMC pregnancy and childbirth. 2019 Dec;19:1-1.
259. Ntabanganyimana E, Giraneza R, Dusabejambo V, Bizimana A, Hamond C, Iyamuremye A, Nshizirungu P, Uzabakiriho R, Munyengabe M, Wunder Jr EA, Page C. Sero-prevalence of anti-Leptospira antibodies and associated risk factors in rural Rwanda: A cross-sectional study. PLoS Neglected Tropical Diseases. 2021 Dec 7;15(12):e0009708.
260. Haule A, Msemwa B, Mgaya E, Masikini P, Kalluvya S. Prevalence of syphilis, neurosyphilis and associated factors in a cross-sectional analysis of HIV infected patients attending Bugando Medical Centre, Mwanza, Tanzania. BMC Public Health. 2020 Dec;20(1):1-2.
261. Taye M, Afework M, Fantaye W, Diro E, Worku A. Congenital anomalies prevalence in Addis Ababa and the Amhara region, Ethiopia: a descriptive cross-sectional study. BMC pediatrics. 2019 Dec;19:1-1.
262. Hrapcak S, Bekele A, Ahmed J, Ayalew J, Gutreuter S, Kumssa H, Antefe T, Mengistu S, Mirkovic K, Dziuban EJ, Ross C. Finding children living with HIV in low-prevalence countries: HIV prevalence and testing yield from 5 entry points in Ethiopia. The Pediatric infectious disease journal. 2021 Nov 8;40(12):1090-5.
263. John W, Mushi V, Tarimo D, Mwingira U. Prevalence and management of filarial lymphoedema and its associated factors in Lindi district, Tanzania: A community-based cross-sectional study. Tropical Medicine & International Health. 2022 Aug;27(8):678-85.
264. Jeremiah K, Lyimo E, Ritz C, PrayGod G, Rutkowski KT, Korsholm KS, Ruhwald M, Tait D, Grewal HM, Faurholt-Jepsen D. Prevalence of Mycobacterium tuberculosis infection as measured by the QuantiFERON-TB Gold assay and ESAT-6 free IGRA among adolescents in Mwanza, Tanzania. Plos one. 2021 Jun 7;16(6):e0252808.
265. Chepkondol GK, Jolly PE, Yatich N, Mbowe O, Jaoko WG. Types and prevalence of HIV-related opportunistic infections/conditions among HIV-positive patients attending Kenyatta National Hospital in Nairobi, Kenya. African Health Sciences. 2020 Jul 22;20(2):615-24.
266. Guyatt H, Muiruri F, Mburu P, Robins A. Prevalence and predictors of underweight and stunting among children under 2 years of age in Eastern Kenya. Public Health Nutrition. 2020 Jun;23(9):1599-608.
267. Tamomh AG, Agena AM, Elamin E, Suliman MA, Elmadani M, Omara AB, Musa SA. Prevalence of cryptosporidiosis among children with diarrhoea under five years admitted to Kosti teaching hospital, Kosti City, Sudan. BMC Infectious Diseases. 2021 Dec;21(1):1-6.
268. Baluku JB, Mayinja E, Mugabe P, Ntabadde K, Olum R, Bongomin F. Prevalence of anaemia and associated factors among people with pulmonary tuberculosis in Uganda. Epidemiology & Infection. 2022;150.
269. Adegbamigbe OJ, Yusuf M, Durowade KA, Oguntoye OO, Ogundare Y. Exposure to patients' sample and prevalence of Hepatitis B and C virus infection among health-care workers in a Nigerian Tertiary Hospital. Annals of African Medicine. 2022 Oct;21(4):322.
270. Joseph Davey DL, Nyemba DC, Gomba Y, Bekker LG, Taleghani S, DiTullio DJ, Shabsovich D, Gorbach PM, Coates TJ, Klausner JD, Myer L. Prevalence and correlates of sexually transmitted infections in pregnancy in HIV-infected and-uninfected women in Cape Town, South Africa. PloS one. 2019 Jul 1;14(7):e0218349.
271. Temesgen MM, Alemu T, Shiferaw B, Legesse S, Zeru T, Haile M, Gelanew T. Prevalence of oncogenic human papillomavirus (HPV 16/18) infection, cervical lesions and its associated factors among women aged 21–49 years in Amhara region, Northern Ethiopia. Plos one. 2021 Mar 24;16(3):e0248949.
272. Cisse M, Sangare I, Djibougou AD, Tahita MC, Gnissi S, Bassinga JK, Konda S, Diallo AH. Prevalence and risk factors of Schistosoma mansoni infection among preschool-aged children from Panamasso village, Burkina Faso. Parasites & Vectors. 2021 Dec;14:1-9.
273. Krings A, Dunyo P, Pesic A, Tetteh S, Hansen B, Gedzah I, Wormenor CM, Amuah JE, Behnke AL, Höfler D, Pawlita M. Characterization of Human Papillomavirus prevalence and risk factors to guide cervical cancer screening in the North Tongu District, Ghana. PLoS One. 2019 Jun 27;14(6):e0218762.

274. Danso F, Appiah MA. Prevalence and associated factors influencing stunting and wasting among children aged 1 to 5 years in Nkwanta South Municipality, Ghana. Nutrition. 2023 Feb 9:111996.
275. Ambaye E, Ormago MD, Ali MM. Sero-prevalence and associated factors of sexually transmitted infections among youth-friendly services Attendees. Plos one. 2023 Jan 23;18(1):e0279900.
276. Coldiron ME, Assao B, Guindo O, Sayinzoga-Makombe N, Koscalova A, Sterk E, Quere M, Ciglenecki I, Mumina A, Atti S, Langendorf C. Prevalence of malaria in an area receiving seasonal malaria chemoprevention in Niger. Malaria Journal. 2021 Dec;20(1):1-8.
277. Akuffo R, Wilson M, Sarfo B, Attram N, Mosore MT, Yeboah C, Cruz I, Ruiz-Postigo JA, Boakye D, Moreno J, Anto F. Prevalence of Leishmania infection in three communities of Oti Region, Ghana. PLoS Neglected Tropical Diseases. 2021 May 27;15(5):e0009413.
278. Cosmas NT, Nimzing L, Egah D, Famooto A, Adebamowo SN, Adebamowo CA. Prevalence of vaginal HPV infection among adolescent and early adult girls in Jos, North-Central Nigeria. BMC Infectious Diseases. 2022 Apr 5;22(1):340.
279. Ashagrie D, Genet C, Abera B. Vancomycin-resistant enterococci and coagulase-negative staphylococci prevalence among patients attending at Felege Hiwot Comprehensive Specialized Hospital, Bahir Dar, Ethiopia. Plos one. 2021 Apr 8;16(4):e0249823.
280. Sassa M, Chadeka EA, Cheruiyot NB, Tanaka M, Moriyasu T, Kaneko S, Njenga SM, Cox SE, Hamano S. Prevalence and risk factors of Schistosoma mansoni infection among children under two years of age in Mbita, Western Kenya. PLoS neglected tropical diseases. 2020 Aug 25;14(8):e0008473.
281. Kagujje M, Somwe P, Hatwiinda S, Bwalya J, Zgambo T, Thornicroft M, Bozzani FM, Moonga C, Muyoyeta M. Cross-sectional assessment of tuberculosis and HIV prevalence in 13 correctional facilities in Zambia. BMJ open. 2021 Sep 1;11(9):e052221.
282. Tuke D, Etu E, Shalemo E. Active trachoma prevalence and related variables among children in a pastoralist community in southern Ethiopia in 2021: a community-based cross-sectional study. The American Journal of Tropical Medicine and Hygiene. 2023 Feb;108(2):252.
283. Ahenkorah B, Nsiah K, Baffoe P, Ofosu W, Gyasi C, Owiredu EW. Parasitic infections among pregnant women at first antenatal care visit in northern Ghana: a study of prevalence and associated factors. PloS one. 2020 Jul 24;15(7):e0236514.
284. Pathirana J, Groome M, Dorfman J, Kwatra G, Boppana S, Cutland C, Jones S, Madhi SA. Prevalence of congenital cytomegalovirus infection and associated risk of in utero human immunodeficiency virus (HIV) acquisition in a high-HIV prevalence setting, South Africa. Clinical Infectious Diseases. 2019 Oct 30;69(10):1789-96.
285. Njoku MO, Iloh KK, Okike CO, Njoku GC, Ojinnaka NC. The prevalence and intensity of intestinal helminths among institutionalized children in three states of South-East Nigeria. Nigerian Journal of Clinical Practice. 2022 May 1;25(5):718-24.
286. Bayih WA, Yitbarek GY, Aynalem YA, Abate BB, Tesfaw A, Ayalew MY, Belay DM, Hailemeskel HS, Alemu AY. Prevalence and associated factors of birth asphyxia among live births at Debre Tabor General Hospital, North Central Ethiopia. BMC pregnancy and childbirth. 2020 Dec;20(1):1-2.
287. Kisindja RM, Tugirimana PL, Prudence MN, Bosunga K, Sihalikyolo JJ, Kayamba PK. Prevalence of gestational diabetes in Eastern Democratic Republic of Congo. BMC Pregnancy and Childbirth. 2022 Dec;22(1):1-6.
288. Nkenfou CN, Fainguem N, Dongmo-Nguefack F, Yatchou LG, Kameni JJ, Elong EL, Samie A, Estrin W, Koki PN, Ndjolo A. Enhanced passive surveillance dengue infection among febrile children: Prevalence, co-infections and associated factors in Cameroon. PLoS Neglected Tropical Diseases. 2021 Apr 16;15(4):e0009316.
289. Mueller A, Fuss A, Ziegler U, Kaatano GM, Mazigo HD. Intestinal schistosomiasis of Ijinga Island, north-western Tanzania: prevalence, intensity of infection, hepatosplenic morbidities and their associated factors. BMC infectious diseases. 2019 Dec;19(1):1-2.
290. Ambe NF, Longdoh NA, Tebid P, Bobga TP, Nkfusai CN, Ngwa SB, Nsai FS, Cumber SN. The prevalence, risk factors and antifungal sensitivity pattern of oral candidiasis in HIV/AIDS patients in Kumba District Hospital, South West Region, Cameroon. Pan African Medical Journal. 2020 May 19;36(1).
291. Okyere B, Owusu-Ofori A, Ansong D, Buxton R, Benson S, Osei-Akoto A, Owiredu EW, Adjei C, Xorse Amuzu E, Marfo Boaheng J, Dickerson T. Point prevalence of asymptomatic Plasmodium infection and the

comparison of microscopy, rapid diagnostic test and nested PCR for the diagnosis of asymptomatic malaria among children under 5 years in Ghana. PLoS One. 2020 Jul 27;15(7):e0232874.
292. Sheehy C, Lawson H, Andriamasy EH, Russell HJ, Reid A, Raderalazasoa GU, Dodge G, Kornitschky R, Penney JM, Ranaivoson TN, Andrianiaina A. Prevalence of intestinal schistosomiasis in pre-school aged children: a pilot survey in Marolambo District, Madagascar. Infectious Diseases of Poverty. 2021 Dec;10(1):1-9.
293. Abraham ZS, Ntunaguzi D, Kahinga AA, Mapondella KB, Massawe ER, Nkuwi EJ, Nkya A. Prevalence and etiological agents for chronic suppurative otitis media in a tertiary hospital in Tanzania. BMC research notes. 2019 Dec;12:1-6.
294. Tigabu A, Taye S, Aynalem M, Adane K. Prevalence and associated factors of intestinal parasitic infections among patients attending Shahura Health Center, Northwest Ethiopia. BMC research notes. 2019 Dec;12:1-8.
295. Muze M, Yesse M, Kedir S, Mustefa A. Prevalence and associated factors of undernutrition among pregnant women visiting ANC clinics in Silte zone, Southern Ethiopia. BMC Pregnancy and Childbirth. 2020 Dec;20:1-8.
296. Lungosi MB, Muzembo BA, Mbendi NC, Nkodila NA, Ngatu NR, Suzuki T, Wada K, Mbendi NS, Ikeda S. Assessing the prevalence of hepatitis B virus infection among health care workers in a referral hospital in Kisantu, Congo DR: A pilot study. Industrial health. 2019;57(5):621-6.
297. Teklu T, Chauhan NM, Lemessa F, Teshome G. Assessment of prevalence of malnutrition and its associated factors among AIDS patients from Asella, Oromia, Ethiopia. BioMed Research International. 2020 Dec 8;2020.
298. Bukar AK, Galadima BG, Zailani SB, Yahaya M, Daggash BB, Yakubu MY, Baba AS, Shettima AB, Kadaura MU. Prevalence of rotavirus isolates in the stools of under-5 children presenting with diarrhoea at University Maiduguri Teaching Hospital, Borno State. Nigerian Journal of Clinical Practice. 2022 Aug 1;25(8):1269-73.
299. Mbunga BK, Mapatano MA, Strand TA, Gjengedal EL, Akilimali PZ, Engebretsen IM. Prevalence of anemia, iron-deficiency anemia, and associated factors among children aged 1–5 years in the rural, malaria-endemic setting of Popokabaka, Democratic Republic of Congo: A cross-sectional study. Nutrients. 2021 Mar 21;13(3):1010.
300. Mohammed H, Oljira L, Roba KT, Ngadaya E, Manyazewal T, Ajeme T, Mnyambwa NP, Fekadu A, Yimer G. Tuberculosis prevalence and predictors among health care-seeking people screened for cough of any duration in Ethiopia: a multicenter cross-sectional study. Frontiers in Public Health. 2022 Feb 25;9:805726.
301. Twesigomwe G, Migisha R, Agaba DC, Owaraganise A, Aheisibwe H, Tibaijuka L, Abesiga L, Ngonzi J, Tornes YF. Prevalence and associated factors of oligohydramnios in pregnancies beyond 36 weeks of gestation at a tertiary hospital in southwestern Uganda. BMC Pregnancy and Childbirth. 2022 Aug 2;22(1):610.
302. Farrant O, Marlais T, Houghton J, Goncalves A, Teixeira da Silva Cassama E, Cabral MG, Nakutum J, Manjuba C, Rodrigues A, Mabey D, Bailey R. Prevalence, risk factors and health consequences of soil-transmitted helminth infection on the Bijagos Islands, Guinea Bissau: A community-wide cross-sectional study. PLoS Neglected Tropical Diseases. 2020 Dec 16;14(12):e0008938.
303. Hayuma PM, Wang CW, Liheluka E, Baraka V, Madebe RA, Minja DT, Misinzo G, Alifrangis M, Lusingu JP. Prevalence of asymptomatic malaria, submicroscopic parasitaemia and anaemia in Korogwe District, north-eastern Tanzania. Malaria Journal. 2021 Dec;20:1-9.
304. Yalew GT, Muthupandian S, Hagos K, Negash L, Venkatraman G, Hagos YM, Meles HN, Weldehaweriat HH, Al-Dahmoshi HO, Saki M. Prevalence of bacterial vaginosis and aerobic vaginitis and their associated risk factors among pregnant women from northern Ethiopia: A cross-sectional study. PloS one. 2022 Feb 25;17(2):e0262692.
305. Yoseph A, Beyene H. The high prevalence of intestinal parasitic infections is associated with stunting among children aged 6–59 months in Boricha Woreda, Southern Ethiopia: a cross-sectional study. BMC Public Health. 2020 Dec;20:1-3.
306. Ojo JA, Adedokun SA, Akindele AA, Olorunfemi AB, Otutu OA, Ojurongbe TA, Thomas BN, Velavan TP, Ojurongbe O. Prevalence of urogenital and intestinal schistosomiasis among school children in South-west Nigeria. PLoS Neglected Tropical Diseases. 2021 Jul 27;15(7):e0009628.

323. Genet A, Dagnew Z, Melkie G, Keleb A, Motbainor A, Mebrat A, Leshargie CT. Prevalence of active trachoma and its associated factors among 1–9 years of age children from model and non-model kebeles in Dangila district, northwest Ethiopia. Plos one. 2022 Jun 15;17(6):e0268441.
324. Aworh MK, Kwaga J, Okolocha E, Mba N, Thakur S. Prevalence and risk factors for multi-drug resistant Escherichia coli among poultry workers in the Federal Capital Territory, Abuja, Nigeria. PloS one. 2019 Nov 21;14(11):e0225379.
325. Tadiwos MB, Kanno GG, Areba AS, Kabthymer RH, Abate ZG, Aregu MB. Sero-prevalence of hepatitis B virus infection and associated factors among pregnant women attending antenatal care services in Gedeo Zone, Southern Ethiopia. Journal of Primary Care & Community Health. 2021 Feb;12:2150132721993628.
326. Kiconco G, Turyasiima M, Ndamira A, Yamile OA, Egesa WI, Ndiwimana M, Maren MB. Prevalence and associated factors of pneumonia among under-fives with acute respiratory symptoms: a cross sectional study at a Teaching Hospital in Bushenyi District, Western Uganda. African Health Sciences. 2021 Dec 14;21(4):1701-0.
327. Kebede BA, Abdo RA, Anshebo AA, Gebremariam BM. Prevalence and predictors of primary postpartum hemorrhage: An implication for designing effective intervention at selected hospitals, Southern Ethiopia. PloS one. 2019 Oct 31;14(10):e0224579.
328. Hechera Y, Dona A. Prevalence of undernutrition and its associated factors among lactating women in the Shebedino District, Sidama Regional State, Ethiopia. INQUIRY: The Journal of Health Care Organization, Provision, and Financing. 2022 Mar 9;59:00469580221087883.
329. Larebo YM, Ermolo NA. Prevalence and risk factors of gestational diabetes mellitus among women attending antenatal care in Hadiya zone public hospitals, southern nation Nationality people region. BioMed Research International. 2021 Apr 5;2021.
330. Manyanga T, Barnes JD, Chaput JP, Dubois L, Katzmarzyk PT, Mire EF, Prista A, Tremblay MS. Prevalence and correlates of objectively measured weight status among urban and rural Mozambican primary schoolchildren: A cross-sectional study. Plos one. 2020 Feb 3;15(2):e0228592.
331. Sandie SM, Sumbele IU, Tasah MM, Kimbi HK. Malaria parasite prevalence and Haematological parameters in HIV seropositive patients attending the regional hospital Limbe, Cameroon: a hospital-based cross-sectional study. BMC Infectious Diseases. 2019 Dec;19:1-1.
332. Kassaw MW, Abebe AM, Tegegne KD, Getu MA, Bihonegn WT. Prevalence and associations of active trachoma among rural preschool children in Wadla district, northern Ethiopia. BMC ophthalmology. 2020 Dec;20:1-0.
333. Geffert K, Maponga TG, Henerico S, Preiser W, Mongella S, Stich A, Kalluvya S, Mueller A, Kasang C. Prevalence of chronic HBV infection in pregnant woman attending antenatal care in a tertiary hospital in Mwanza, Tanzania: a cross-sectional study. BMC infectious diseases. 2020 Dec;20(1):1-0.
334. Getaneh DK, Hordofa LO, Ayana DA, Tessema TS, Regassa LD. Prevalence of Escherichia coli O157: H7 and associated factors in under-five children in Eastern Ethiopia. Plos one. 2021 Jan 28;16(1):e0246024.
335. Roberts JS, Hahn EA, Black J, Maharaj R, Farley JE, Redd AD, Reynolds SJ, Quinn TC, Hansoti B. Determining the prevalence of tuberculosis in emergency departments in the Eastern Cape region of South Africa and the utility of the World Health Organization tuberculosis screening tool. South African Medical Journal. 2021;111(9):872-8.
336. Mekonnen J, Kassim J, Ahmed M, Gebeyehu N. Prevalence of active trachoma and associated factors among children 1–9 years old at Arsi Negele Town, West Arsi Zone, Oromia Regional State, Southern Ethiopia. Plos one. 2022 Oct 7;17(10):e0273808.
337. Kassie BA, Yenus H, Berhe R, Kassahun EA. Prevalence of sexually transmitted infections and associated factors among the University of Gondar students, Northwest Ethiopia: a cross-sectional study. Reproductive health. 2019 Dec;16:1-8.
338. Aza'ah RA, Sumo L, Ntonifor NH, Bopda J, Bamou RH, Nana-Djeunga HC. Point prevalence mapping reveals hotspot for onchocerciasis transmission in the Ndikinimeki Health District, Centre Region, Cameroon. Parasites & Vectors. 2020 Dec;13(1):1-8.
339. Tadesse AD, Anto TG, Birhanu MY, Agedew E, Yimer B, Abejie AN. Prevalence of undernutrition and its associated factors among older adults using Mini Nutritional Assessment tool in Womberma district, West Gojjam Zone, Amhara Region, North West Ethiopia, 2020. Plos one. 2023 Feb 24;18(2):e0274557.

357. Tsegaye Sahle E, Blumenthal J, Jain S, Sun S, Young J, Manyazewal T, Woldeamanuel H, Teferra L, Feleke B, Vandenberg O, Rey Z. Bacteriologically-confirmed pulmonary tuberculosis in an Ethiopian prison: prevalence from screening of entrant and resident prisoners. PloS one. 2019 Dec 12;14(12):e0226160.
358. Mubiru IS, Kasirye PG, Hume H, Ndeezi G. Prevalence and factors associated with Helicobacter Pylori infection among children with sickle cell anemia attending Mulago hospital, in Uganda. African Health Sciences. 2022 Jul 29;22(2):135-45.
359. Imboumy-Limoukou RK, Maghendji-Nzondo S, Ondo-Enguier PN, Niemczura De Carvalho J, Tsafack-Tegomo NP, Buekens J, Okouga AP, Mouinga-Ondeme A, Kwedy Nolna S, Lekana-Douki JB. Malaria in children and women of childbearing age: infection prevalence, knowledge and use of malaria prevention tools in the province of Nyanga, Gabon. Malaria journal. 2020 Dec;19(1):1-8.
360. Patel NH, Meier-Stephenson V, Genetu M, Damtie D, Abate E, Alemu S, Aleka Y, Van Marle G, Fonseca K, Coffin CS, Deressa T. Prevalence and genetic variability of occult hepatitis B virus in a human immunodeficiency virus positive patient cohort in Gondar, Ethiopia. PLoS One. 2020 Nov 19;15(11):e0242577.
361. Assefa G, Alemu M, Ayehu A. High Prevalence of Hookworm Species and Associated Factors among Soil-Transmitted Helminth-Infected Household Contacts in Burie Zuria District, Northwest Ethiopia: A Community-Based Cross-Sectional Study. BioMed Research International. 2023 Jan 7;2023.
362. Yakubu A, Hali B, Maiyaki AS. Prevalence and risk factors for hepatitis c virus co-infection among human immunodeficiency virus-infected patients and effect of hepatitis c virus infection on acquired immunodeficiency syndrome cases at baseline. Annals of African Medicine. 2021 Oct;20(4):297.
363. Mebratu W, Wedajo S, Mohammed S, Endawkie A, Damtew Y. Prevalence and associated factors of tuberculosis among isoniazid users and non-users of HIV patients in Dessie, Ethiopia. Scientific Reports. 2022 Aug 5;12(1):13500.
